# Estimating and predicting kidney function decline in the general population

**DOI:** 10.1101/2024.03.30.24305107

**Authors:** Masao Iwagami, Kazunori Odani, Tomoki Saito

## Abstract

**Introduction:** We aimed to estimate the rate of kidney function decline over 10 years in the general population and develop a machine learning model to predict it.

**Methods:** We used the JMDC database from 2012 to 2021, which includes company employees and their family members in Japan, where annual health checks are mandated for people aged 40–74 years. We estimated the slope (average change) of estimated glomerular filtration rate (eGFR) over a period of 10 years. Then, using the annual health-check results and prescription claims for the first five years from 2012 to 2016 as predictor variables, we developed an XGBoost model, evaluated its prediction performance with the root mean squared error (RMSE), R^2^, and area under the receiver operating characteristic curve (AUROC) for rapid decliners (defined as the slope <-3 ml/min/1.73 m^2^/year) using 5-fold cross validation, and compared these indicators with those of the linear regression model using only eGFR data from 2012 to 2016.

**Results:** We included 126 424 individuals (mean age, 45.2 years; male, 82.4%; mean eGFR, 79.0 ml/min/1.73 m^2^ in 2016). The mean slope was -0.89 (standard deviation, 0.96) ml/min/1.73 m^2^/year. The predictive performance of the XGBoost model (RMSE, 0.78; R^2^, 0.35; and AUROC, 0.89) was better than that of the linear regression model using only eGFR data (RMSE, 1.94; R^2^, -3.03; and AUROC, 0.79).

**Conclusion:** Application of machine learning to annual health-check and claims data could predict the rate of kidney function decline, whereas the linear regression model using only eGFR data did not work.

## Introduction

Chronic kidney disease (CKD) is a large burden in the society as it is associated with increased risk of cardiovascular and non-cardiovascular diseases, as well as the health care costs, especially if patients require renal replacement therapy (RRT).^1–4^ Most people in the general population have normal kidney function (i.e., glomerular filtration rate [GFR]) at birth, whereas the GFR naturally decreases with age, with faster decline among people with risk factors such as diabetes.^5^ An old study estimated that the rate of GFR decline was -0.75 ml/min/year in the generally healthy population.^6^ Since then, there have been a number of studies estimating the rate of kidney function decline,^7^ but their study periods are often short and only a few studies targeting the general population without CKD are reported.

A number of clinical trials and observational studies have set the study endpoints as the time to dialysis initiation or the time to a 30% or 40% drop in estimated GFR (eGFR), mostly among patients at high risk for these events, such as those with late stage CKD.^8,9^ However, the incidence of these outcomes is low in the early stage of CKD or in the general population.^9^ Meanwhile, the rate of kidney function decline or slope (average change) of eGFR can be calculated for individuals and could be a surrogate endpoint for clinical trials, even in the early stage of CKD and in the general population.^10–14^

In observational studies, prediction models have been developed for the time to dialysis initiation^15–17^ or the time to a 30% or 40% drop in eGFR,^18–20^ showing good discrimination ability and/or calibration. However, to the best of our knowledge, no previous study has developed a prediction model for the rate of kidney function decline as a continuous variable. Such prediction model would be useful for stratifying the general population and identifying those with rapid decline in kidney function. To date, no consensus has been reached on the definition of rapid decliners,^7^ and therefore, the prediction of a continuous (rather than a dichotomous) outcome would have a wider application.

In Japan, the government introduced a specific health checkup system in 2008, which obliges all insurers to provide annual health checkups for insured persons aged 40–74 years.^21^ Notably, under employee insurance, the attendance rates for annual checkups are high, approximately >80% (>90% among men) among company employees.^22^ Utilizing this situation, we aimed (i) to estimate the rate of kidney function decline in a period of 10 years using data obtained from the JMDC database, a large database of large and middle-scale companies and their family members in Japan and (ii) to develop a prediction model based on annual health checkup data and claims for the first five years. Machine learning has been used to handle a large number of candidate predictor variables and their potential interactions.

## Methods

### Data source

The details of the JMDC database have been described elsewhere.^23^ In brief, the JMDC database was developed by the JMDC Co. This database is a large-scale database covering Japanese health insurance union members, including employees of large- and middle-scale companies and their family members aged <75 years; it includes all claims for outpatient treatment, hospitalization, and prescriptions and dispensations of drugs, as well as the results of annual health checkups. Annual health checkups are required by law for insured persons aged 40–74 years,^21^ whereas those aged <40 years can also undergo annual checkups. Annual health checkups are usually conducted in the facilities of health insurance unions with which the companies are affiliated. The details of the annual health checkups are listed in the “Predictor variables” section below. Serum creatinine measurement is optional but depends on the decision of each health insurance union rather than on the medical conditions of the participants. For the present study, we used the most recent 10-year data from April 2012 to March 2022 (i.e., from 2012 to 2021 financial years).

The data used in this study were anonymized and processed anonymously by JMDC, Inc. This study was approved by the Ethics Committee of The Research Institute of Healthcare Data Science (Date of approval, October 30, 2023; Approval number, RI 2023003).

### Study population

First, in the JMDC database, we identified people with annual health checkup results (including serum creatinine) for five consecutive years, from 2012 to 2016. We excluded patients receiving RRT (identified as Japanese procedure codes J038 for hemodialysis, J042 for peritoneal dialysis, and K780 for kidney transplantation) from 2012 to 2016 or those with an eGFR <15 ml/min/1.73 m^2^ in 2016. Among the remaining individuals, we further identified those with annual health checkup results (including serum creatinine) for the latter five consecutive years, from 2017 to 2021. We identified and excluded patients who underwent RRT between 2017 and 2021 because their serum creatinine levels did not reflect their GFRs.

Consequently, the study population consisted of people with annual health checkup results (including serum creatinine) for 10 consecutive years, from 2012 to 2021, who did not receive RRT.

### Outcome definition

The outcome of interest was the slope (average change) of eGFR during the 10 years from 2012 to 2021, which was estimated using unadjusted linear regression. eGFR was calculated using the following Japanese estimation formula^24^:

eGFR=194×Cr^(−1.094)×Age^(−0.287) (×0.739 for women)

### Predictor variables

We used the annual health checkup results for the first five years, from 2012 to 2016. As demonstrated prior,^23^ the mandatory annual health checkups in Japan generally include both objective and subjective (self-reported) findings. Objective findings include body mass index (BMI), abdominal circumference, systolic blood pressure (sBP), diastolic blood pressure (dBP), triglyceride (TG), high density lipoprotein (HDL) cholesterol, low density lipoprotein (LDL) cholesterol, total cholesterol, aspartate aminotransferase (ALT), alanine aminotransferase (ALT), gamma glutamyl transpeptidase (γ-GTP), fasting blood sugar, casual blood sugar, hemoglobin A1c (HbA1c) according to the National Glycohemoglobin Standardization Program, hematocrit, hemoglobin content, erythrocyte count, serum uric acid, urinary sugar (dipstick test), and uric protein (dipstick test). Among subjective (self-reported) findings,^23^ we used the information pertaining to current smoking status (yes or no), drinking habits (every day, sometimes, or rarely/none), and exercise habit (yes or no for ≥2 times/week for ≥30 min in the past year).

In addition, using the prescription records in the medical claims, we identified the presence or absence in the use of lipid-lowering agents (any), statins, antidiabetic drugs (any), sodium-glucose transport protein 2 (SGLT2) inhibitors, antiplatelet drugs, antihypertensive drugs (any), and angiotensin converting enzyme inhibitors (ACEI) or angiotensin II receptor blockers (ARB), which are recorded as the Anatomical Therapeutic Chemical (ATC) Classification codes (**Supplementary Table S1**).

**Table 1** displays the list of predictor variables and their distributions (mean and standard deviation [SD] for continuous variables and number and percentage for categorical variables) in 2016, whereas **Supplementary Table S2** shows all predictor variables from 2012 to 2016 that were used for prediction. In addition, the slope (average change) of eGFR during the five years was used for prediction.

**Table 1.**
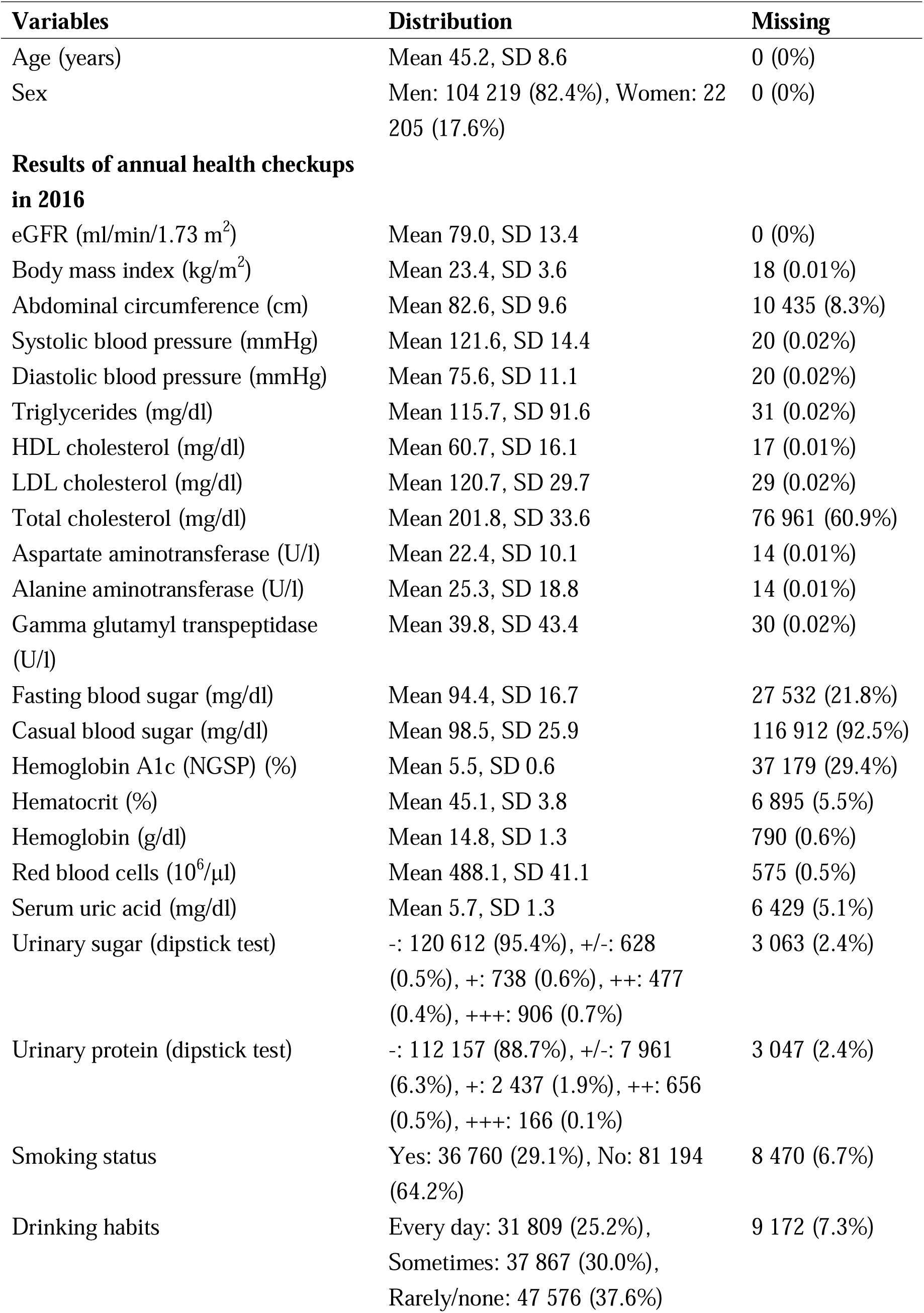
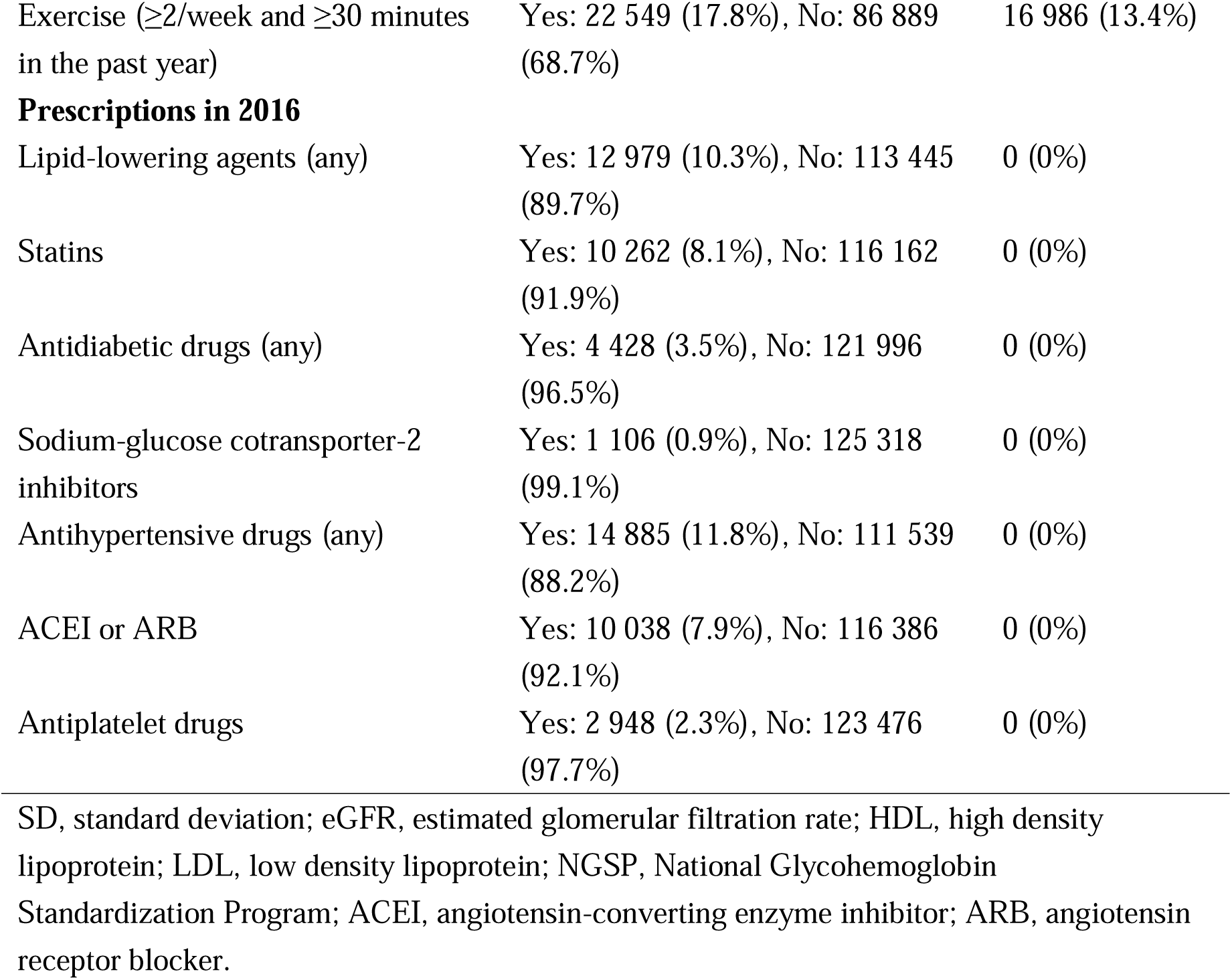
List of predictor variables in 2016 and distributions (n=126 424)

| Variables | Distribution | Missing |
| --- | --- | --- |
| Age (years) | Mean 45.2, SD 8.6 | 0 (0%) |
| Sex | Men: 104 219 (82.4%), Women: 22 205 (17.6%) | 0 (0%) |
| **Results of annual health checkups in 2016** |  |  |
| eGFR (ml/min/1.73 m<sup>2</sup>) | Mean 79.0, SD 13.4 | 0 (0%) |
| Body mass index (kg/m<sup>2</sup>) | Mean 23.4, SD 3.6 | 18 (0.01%) |
| Abdominal circumference (cm) | Mean 82.6, SD 9.6 | 10 435 (8.3%) |
| Systolic blood pressure (mmHg) | Mean 121.6, SD 14.4 | 20 (0.02%) |
| Diastolic blood pressure (mmHg) | Mean 75.6, SD 11.1 | 20 (0.02%) |
| Triglycerides (mg/dl) | Mean 115.7, SD 91.6 | 31 (0.02%) |
| HDL cholesterol (mg/dl) | Mean 60.7, SD 16.1 | 17 (0.01%) |
| LDL cholesterol (mg/dl) | Mean 120.7, SD 29.7 | 29 (0.02%) |
| Total cholesterol (mg/dl) | Mean 201.8, SD 33.6 | 76 961 (60.9%) |
| Aspartate aminotransferase (U/l) | Mean 22.4, SD 10.1 | 14 (0.01%) |
| Alanine aminotransferase (U/l) | Mean 25.3, SD 18.8 | 14 (0.01%) |
| Gamma glutamyl transpeptidase (U/l) | Mean 39.8, SD 43.4 | 30 (0.02%) |
| Fasting blood sugar (mg/dl) | Mean 94.4, SD 16.7 | 27 532 (21.8%) |
| Casual blood sugar (mg/dl) | Mean 98.5, SD 25.9 | 116 912 (92.5%) |
| Hemoglobin A1c (NGSP) (%) | Mean 5.5, SD 0.6 | 37 179 (29.4%) |
| Hematocrit (%) | Mean 45.1, SD 3.8 | 6 895 (5.5%) |
| Hemoglobin (g/dl) | Mean 14.8, SD 1.3 | 790 (0.6%) |
| Red blood cells (10<sup>6</sup>/μl) | Mean 488.1, SD 41.1 | 575 (0.5%) |
| Serum uric acid (mg/dl) | Mean 5.7, SD 1.3 | 6 429 (5.1%) |
| Urinary sugar (dipstick test) | -: 120 612 (95.4%), +/-: 628 (0.5%), +: 738 (0.6%), ++: 477 (0.4%), +++: 906 (0.7%) | 3 063 (2.4%) |
| Urinary protein (dipstick test) | -: 112 157 (88.7%), +/-: 7 961 (6.3%), +: 2 437 (1.9%), ++: 656 (0.5%), +++: 166 (0.1%) | 3 047 (2.4%) |
| Smoking status | Yes: 36 760 (29.1%), No: 81 194 (64.2%) | 8 470 (6.7%) |
| Drinking habits | Every day: 31 809 (25.2%), Sometimes: 37 867 (30.0%), Rarely/none: 47 576 (37.6%) | 9 172 (7.3%) |
| Exercise ($\geq 2$/week and $\geq 30$ minutes in the past year) | Yes: 22 549 (17.8%), No: 86 889 (68.7%) | 16 986 (13.4%) |
| **Prescriptions in 2016** |  |  |
| Lipid-lowering agents (any) | Yes: 12 979 (10.3%), No: 113 445 (89.7%) | 0 (0%) |
| Statins | Yes: 10 262 (8.1%), No: 116 162 (91.9%) | 0 (0%) |
| Antidiabetic drugs (any) | Yes: 4 428 (3.5%), No: 121 996 (96.5%) | 0 (0%) |
| Sodium-glucose cotransporter-2 inhibitors | Yes: 1 106 (0.9%), No: 125 318 (99.1%) | 0 (0%) |
| Antihypertensive drugs (any) | Yes: 14 885 (11.8%), No: 111 539 (88.2%) | 0 (0%) |
| ACEI or ARB | Yes: 10 038 (7.9%), No: 116 386 (92.1%) | 0 (0%) |
| Antiplatelet drugs | Yes: 2 948 (2.3%), No: 123 476 (97.7%) | 0 (0%) |

SD, standard deviation; eGFR, estimated glomerular filtration rate; HDL, high density lipoprotein; LDL, low density lipoprotein; NGSP, National Glycohemoglobin Standardization Program; ACEI, angiotensin-converting enzyme inhibitor; ARB, angiotensin receptor blocker.

### Statistical analysis

First, we showed the distribution of outcome variable (i.e., the slope of eGFR during the 10 years) and estimated the mean and standard deviation, overall and by age group (<40, 40–49, 50–59, and ≥60 years), sex, and Kidney Disease Improving Global Outcomes (KDIGO) GFR stages (eGFR ≥90, 60–89, 45–59, 30–44, and 15–29 ml/min/1.73 m^2^) in 2016.

For the model development, we used the XGBoost regression model^25^, because it is generally known to show high predictive performance in the case of table data. The implementation was based on the “xgboost” package (version: 1.7.5) of Python. For the hyperparameters, *eta* (step size shrinkage used in the update to prevent overfitting) was set to 0.05, *subsample* (i.e., subsample ratio of the training instances) was set to 0.9, and *colsample_bytree* (i.e., subsampling of columns) was set to 0.8. With a grid search, the *max_depth* (i.e., maximum depth of a tree) and *min_child_weight* (i.e., minimum sum of instance weights [Hessian] needed in a child) were set to 4 and 16, respectively. *n_estimators* was set by early stopping. The other hyperparameters were set to be default values (“XGBoost-link”). We input the annual health checkup data and prescription data for 2016 into the model as they are. For data from 2012 to 2015, we input the subjective (self-reported) findings and prescription data as they are, whereas we calculated and used the difference in values of objective findings between each year and 2016 for each individual. Missing values were input into the XGBoost model as they are.

For model validation, using the 5-fold cross validation, we evaluated the root mean squared error (RMSE) and R^2^ as the prediction performance:

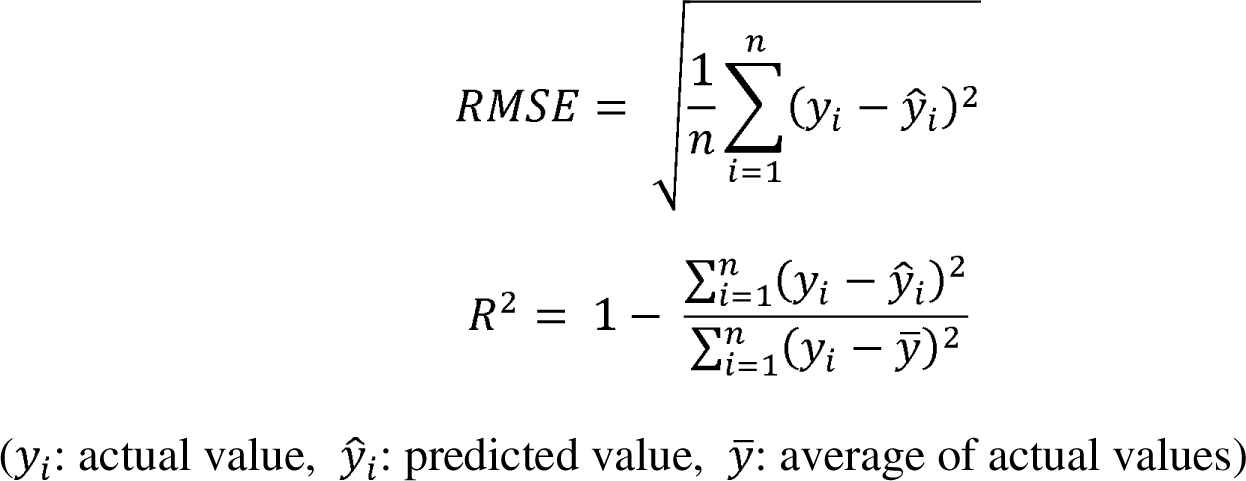

In addition, considering that the identification of rapid decliners (defined in the present study as the slope <-3 ml/min/1.73 m^2^/year^14^) is clinically important, we estimated the area under the receiver operating characteristic curve (AUROC) for the rapid decliners using 5-fold cross validation. For cross-validation, we merged the validation data of each fold to estimate the RMSE, R^2^, and AUROC of the entire training dataset.

For comparison, we also estimated these indicators (i.e., RMSE, R^2^, and AUROC) in a crude linear regression model based only on the eGFR values from 2012 to 2016. We aimed to understand the extent to which our XGBoost model was superior to the simplest linear regression model.

To interpret the model, we used the Shapley Addictive explanation (SHAP) function in the XGBoost model.^26^ We estimated the SHAP feature importance, which is the mean of the absolute SHAP values (i.e., the contribution of each feature to the outcome). For the predictor variables with higher feature importance, we depicted SHAP dependence plots to determine the impact of the increase or decrease in each feature on the predicted value.

All data management and statistical analyses were performed using Python (version 3.8.10).

## Results

In the JMDC database, we identified 183 485 people who underwent annual health checkups for five consecutive years from 2012 to 2016 (**Supplementary Figure S1**). We then excluded 107 patients who underwent RRT and 28 patients with eGFR <15 ml/min/1.73 m^2^. Of the remaining 183 350 people, we excluded 56 926 without annual health checkup results (including serum creatinine) for five consecutive years from 2017 to 2021, including 109 patients who received RRT during this period. The remaining 126 424 individuals with consecutive measurements of serum creatinine levels from 2012 to 2021 were included in the subsequent analyses. A comparison of the predictor variables between the two groups (i.e., 126 424 and 56 926 people with and without annual health checkup results from 2017 to 2021) suggested that older people and women were more likely to retire and lose employee insurance to quit from the JMDC database, whereas the annual health checkup results from 2012 to 2016 were not very different between the two groups (**Supplementary Table S2**).

The mean age of the 126 424 people was 45.2 (SD, 8.6) years, 104 219 (82.4%) were male, and the mean eGFR was 79.0 (SD, 13.4) ml/min/1.73 m^2^. According to the KDIGO GFR categories, 23 850 (18.9%), 95 516 (75.6%), 6 781 (5.4%), 247 (0.2%), and 30 (0.02%) accounted for stages G1, G2, G3a, G3b, and G4 (eGFR ≥90, 60–89, 45–59, 30–44, and 15–29 ml/min/1.73 m^2^), respectively.

Figure 1 shows the distribution of eGFR slope for the 10 years, with a mean value of -0.89 (SD, 0.96) ml/min/1.73 m^2^/year. Among the 126 424 people, 2 511 (2.0%) were rapid decliners, with <-3 ml/min/1.73 m^2^/year. The distributions of eGFR slope by age group, sex, and KDIGO GFR stage are shown in **Supplementary Table S3**.

**Figure 1.**
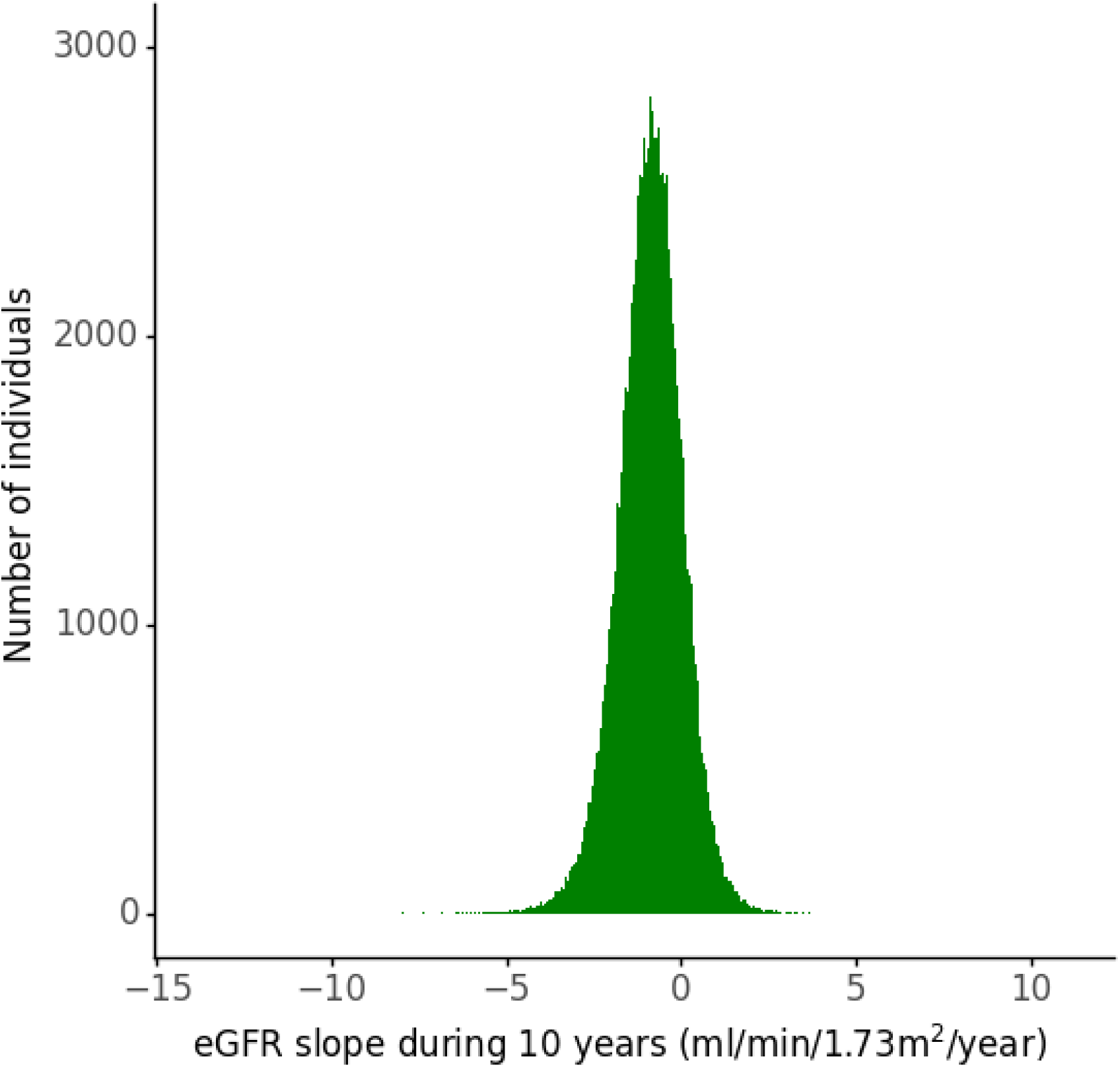
Distribution of the slope (average change) of kidney function decline over 10 years among the study participants (n=126 464) eGFR = estimated glomerular filtration rate.

Figure 2 shows scatter plots of the predicted and actual values in each model, suggesting that the prediction was better in the developed XGBoost model than in the linear regression model using only eGFR data from 2012 to 2016. The RMSE and R^2^ of the developed XGBoost model were 0.78 and 0.35, respectively, while those of the linear regression model using only eGFR data were 1.94 and -3.03, respectively. To discriminate the rapid decliners, the AUROC of the XGBoost model was 0.89, whereas that of the linear regression model using only eGFR data was 0.79. The ROC curves are shown in Figure 3.

**Figure 2.**
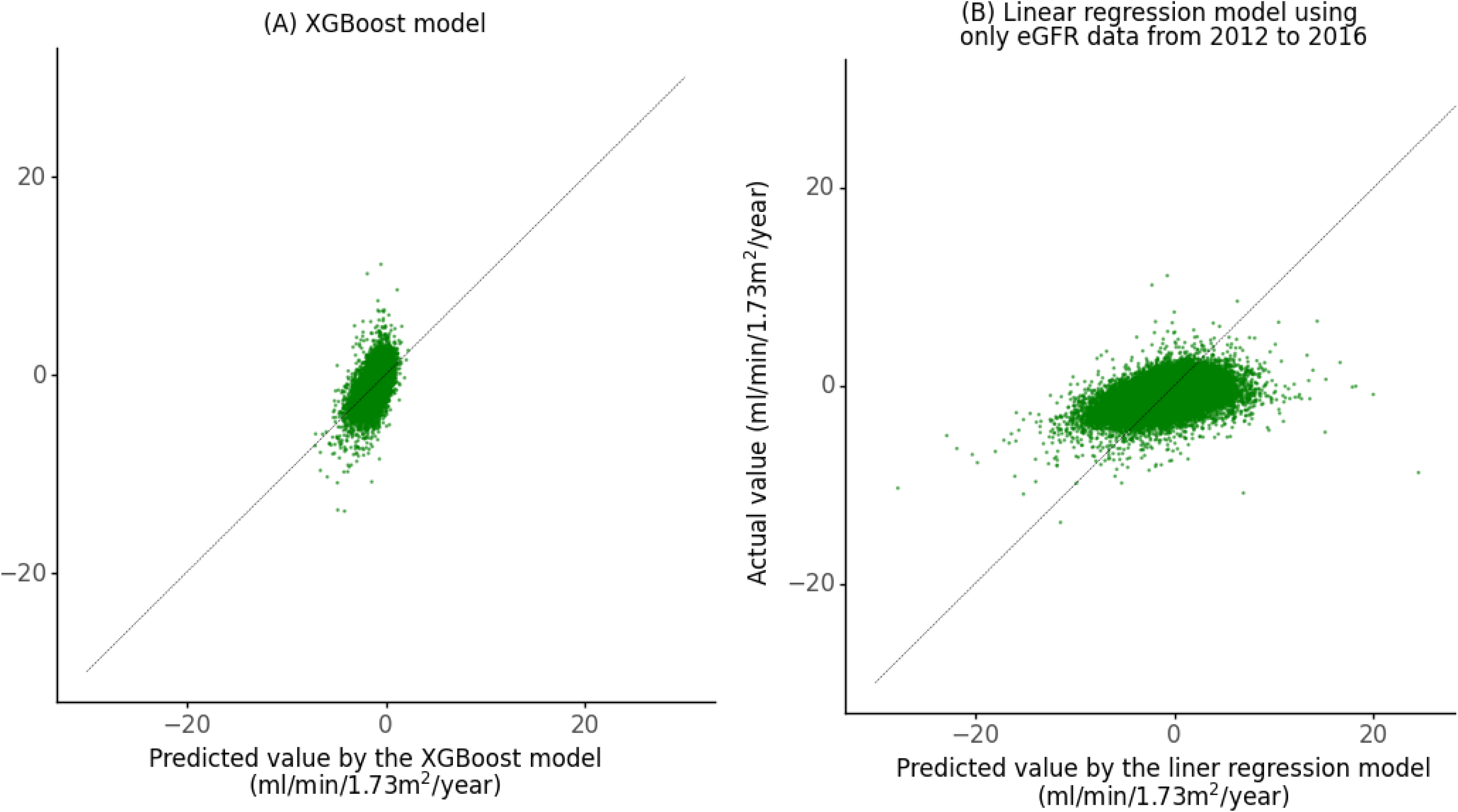
Scatter plots of predicted and actual values (A) in the XGBoost model and (B) in the linear regression model using only eGFR data from 2012 to 2016. eGFR = estimated glomerular filtration rate.

**Figure 3.**
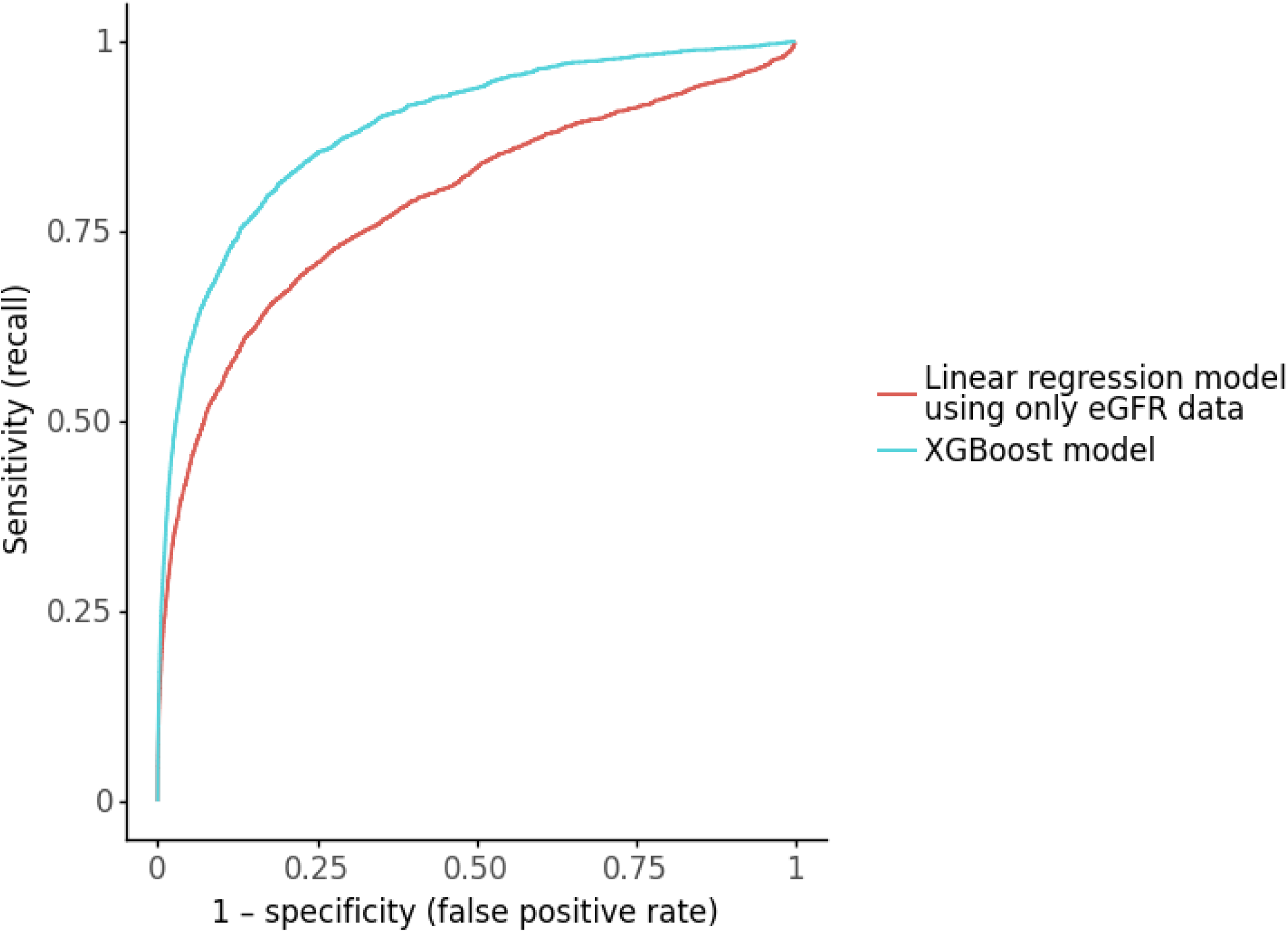
Receiver operating characteristic curves for the rapid decliners (<-3 ml/min/1.73 m^2^/year) eGFR = estimated glomerular filtration rate.

Figure 4 shows the ranking of the predictor variables with higher feature importance in the developed XGBoost model. The eGFR-related features, particularly the eGFR slope from 2012 to 2016 and the eGFR in 2016, ranked high. Several parameters measured in the annual health checkups, including hematocrit, HbA1c, γ-GTP, BMI, HDL cholesterol, serum uric acid, and dBP, were also ranked. Figure 5 shows the SHAP dependence plots of the selected predictor variables with more important features (the difference in values between 2012–2015 and 2016 is not shown because its interpretation was difficult). The model learned to reduce the predicted value (i.e., slope of eGFR for the 10 years) when the slope in the first five years was smaller, and when the eGFR in 2016 was higher in the range above approximately 60 ml/min/1.73 m^2^. The figures also suggest negative associations with Hb1Ac (in the range of above approximately 6.5%), serum uric acid, and dBP (in the range of above approximately 80 mmHg) and positive associations with hematocrit, γ-GTP, and HDL cholesterol. There was a U-shaped association with BMI. Men were more likely to show lower predictive values than women.

**Figure 4.**
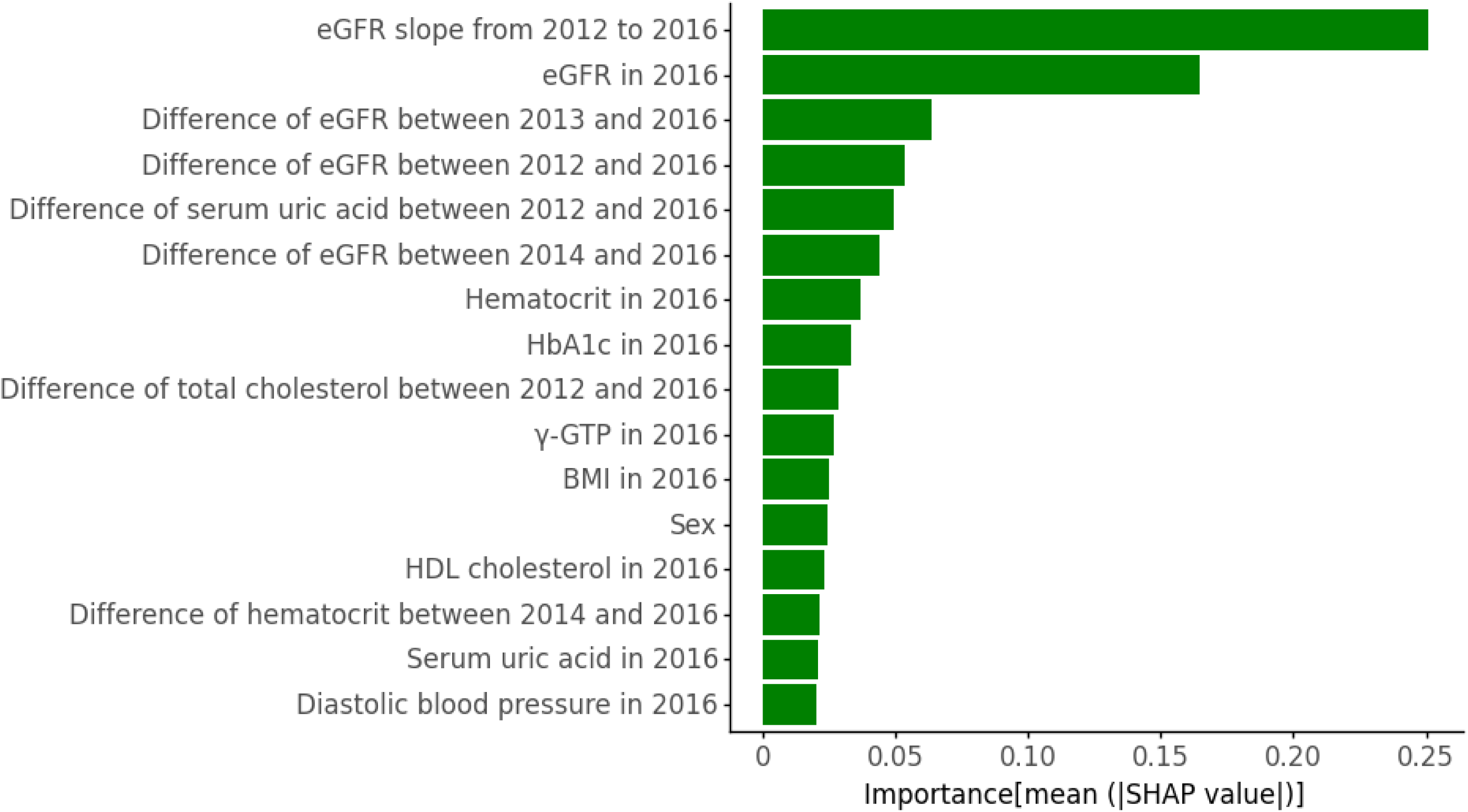
Ranking of predictor variables with higher feature importance in the XGBoost model. eGFR = estimated glomerular filtration rate, HbA1c = hemoglobin A1c, γ-GTP = gamma glutamyl transpeptidase, BMI = body mass index, HDL = high density lipoprotein.

**Figure 5.**
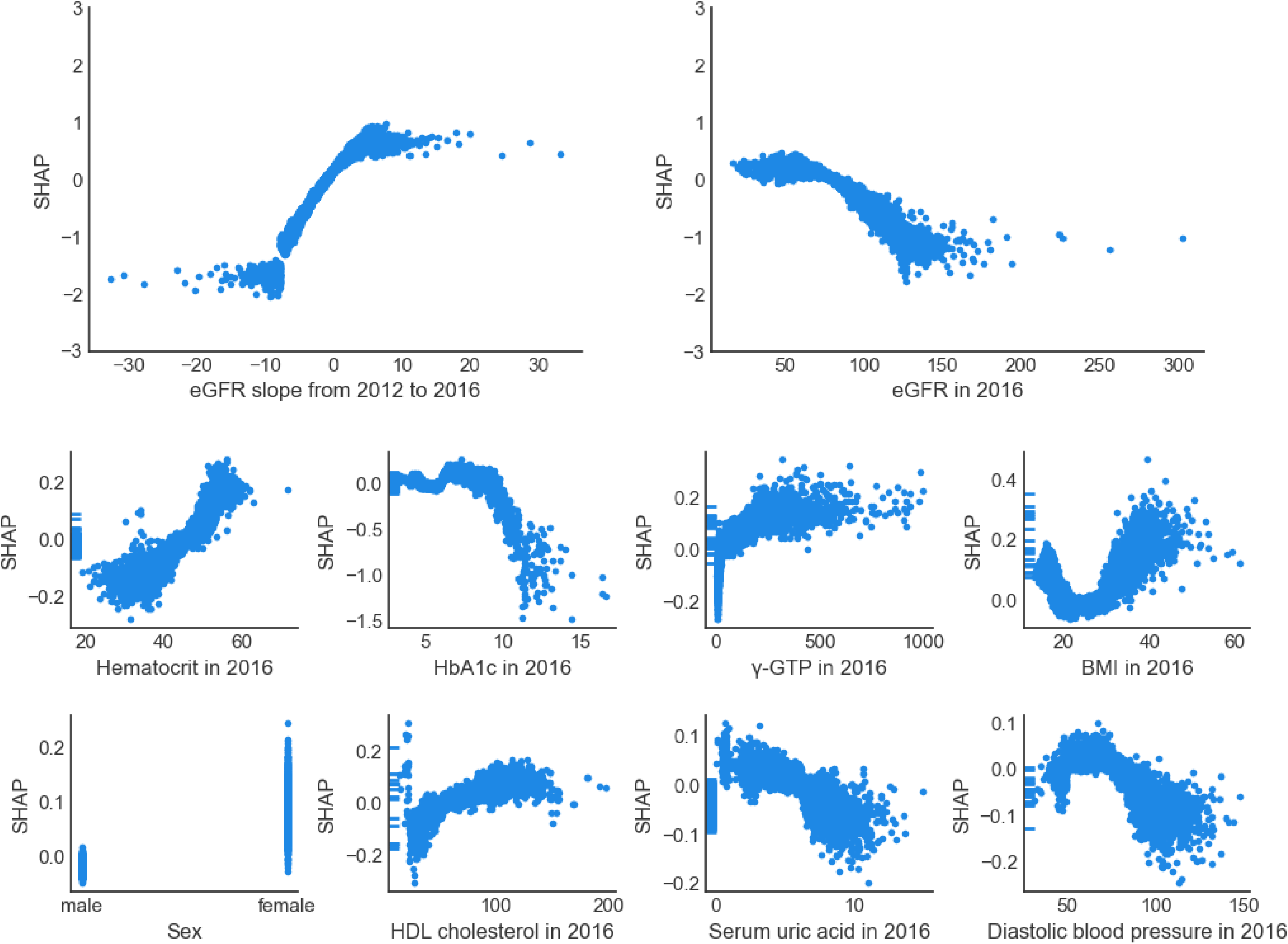
Shapley Addictive explanation (SHAP) dependence plots of predictor variables with higher important features in the XGBoost model. eGFR = estimated glomerular filtration rate, HbA1c = hemoglobin A1c, γ-GTP = gamma glutamyl transpeptidase, BMI = body mass index, HDL = high density lipoprotein.

## Discussion

Using the JMDC claims database covering the general population, we estimated the rate of kidney function decline over 10 years and developed a machine learning model (XGBoost model) to predict this decline based on annual health checkups and prescription data for the first five years. The predictive performance of the model was good or moderate (RMSE, 0.78; R^2^, 0.35; and AUROC for the rapid decliners, 0.89), whereas the linear regression model using only eGFR values did not work (RMSE, 1.94; R^2^, -3.03; and AUROC for the rapid decliners, 0.79). The top features of the developed model were dominated by eGFR-related features, whereas known risk factors for kidney function decline, such as HbA1c, contributed to the prediction.

Among the study population consisting of large- or medium-sized company employees and their family members, most of whom had normal range of kidney function, the mean slope of eGFR for the 10 years was -0.89 (SD, 0.96) ml/min/1.73 m^2^/year. This was similar to an old US study estimating the mean decrease in creatinine clearance to be -0.75 ml/min/year among normal volunteers.^6^ In a recent study conducted in a single health checkup center of Japan, excluding patients with any comorbidities, the mean eGFR decline rate was -1.07 (SD 0.42) ml/min/1.73 m^2^/year.^27^ Those with higher eGFR at baseline were generally more likely to show faster decline, which is in line with the present study (**Supplementary Table S3**).

To the best of our knowledge, this is the first study to predict the rate of kidney function decline as a continuous variable. First, we found it difficult to predict the rate of kidney function decline over 10 years from only eGFR values for the first five years, with an R^2^ of -3.03, which is even worse than a prediction assuming the average value for all individuals (i.e., R^2^ of 0). In other words, the correlation between the slope for the 10 years and that for the first five years was weak. This may be partly due to fluctuations in measured serum creatinine over time, although mandatory annual health checkups in Japanese companies are usually conducted at the same place every year and when participants are not sick. Another reason could be that the rate of kidney function decline is also affected by patient demographics; known risk factors for CKD or ESRD, such as diabetes and hypertension; lifestyle factors, such as smoking and exercise; and drugs protecting the kidneys, such as ACEI/ARB and SGLT2 inhibitors. Therefore, we additionally used these variables as predictors and observed a large increase in R^2^ to 0.35. However, the predictive ability was far from perfect (R^2^ of 1.00). The remaining possibility is that there may be measurement errors in predictor variables or unknown factors predicting the rate of kidney function decline. Meanwhile, the discrimination ability of the XGBoost model for the rapid decliners (eGFR slope <-3 ml/min/1.73 m^2^/year) was seemingly very good, with an AUROC of 0.89.

The feature importance and SHAP dependence plots in the established XGBoost model were remarkable. First, the coefficient (slope) of eGFR for the first five years correlated with the outcome in the same direction, which is intuitive. Meanwhile, the absolute value of eGFR in 2016 was negatively correlated with the outcome in the range above 60 ml/min/1.73 m^2^, meaning that participants with higher eGFR were more likely (i.e., participants with lower eGFR were less likely) to show faster kidney function decline. This phenomenon was also suggested in a previous Japanese study,^27^ wherein the authors speculated that a compensatory mechanism might work as kidney function decreases. For features other than eGFR-related features, negative associations with Hb1Ac, serum uric acid, and dBP, as well as positive associations with hematocrit and HDL cholesterol, have been suggested in some previous studies.^27–32^ Meanwhile, a U-shaped association with BMI, suggesting that those with a normal BMI range were most likely to show a faster decline, may conflict with previous overseas studies suggesting that obesity is a risk factor for ESRD.^33,34^ Further studies are warranted to examine whether this finding is specific to the prediction of the rate of kidney function decline in the general population or whether this is ascribed to the difficulty in estimating accurate eGFR from the existing formula^24^ in obese patients.

The strength of this study is that we used consecutive 10-year annual health checkup data for the general adult population. A systematic review of kidney disease progression^7^ indicated that following the same population for a long time is practically difficult, and only one study was reported to achieve the mean follow-up of over 10 years.^35^ The mandatory health checkup system for people aged 40–74 years in Japan made the present study feasible.

However, this study has some limitations. The database consists of large- and medium-sized company employees and their family members; therefore, their socioeconomic status is expected to be higher than the average in Japan. Accordingly, their health-related behaviors (e.g., smoking and drinking) may be better than those of other Japanese citizens, whereas they may be exposed to stress specific to company employees (e.g., sedentary lifestyles). Therefore, it is unknown whether and to what extent the findings of the present study can be generalized to other citizens in Japan as well as to those living in foreign countries. Second, loss to follow-up could cause selection bias, especially if unhealthy people are more likely to be lost to follow-up.^7^ In the present study, older people and women were more likely to quit the JMDC database. We speculate that the main reasons for the loss to follow-up were social (e.g., retirement at age 60–65, retirement due to pregnancy and childbirth) and not directly associated with the health status of study participants, but we could not confirm the exact reasons. Thus, the effect of the loss to follow-up on our study results is unknown, although the follow-up rate in the present study is better than those in previous studies with long follow-up periods.^27–32^ Third, we obtained the annual health checkup results from health insurance associations instead of the laboratories to measure blood samples, including serum creatinine. Although we believe that creatinine was measured using an internationally standardized enzymatic method (traceable to isotope dilution mass spectrometry) during the study period, creatinine measurements might not be perfectly standardized across laboratories in Japan. However, the influence of this issue in estimating the rate of kidney function decline seems to be small because blood samples from the same individual are expected to be sent to the same laboratories every year. Finally, as discussed above, there may be measurement errors in some predictor variables, especially in self-reported variables such as smoking, drinking, and exercise habits. Furthermore, there may be unknown, and therefore, unmeasured factors predicting the rate of kidney function decline. Further studies are warranted to identify novel risk factors for the rapid decline in kidney function and reassess the performance of the prediction model.

In conclusion, using a large database of company employees and their family members in Japan, we estimated the rate of kidney function decline over 10 years and developed a machine learning prediction model based on annual health checkup results and claims for the first five years. The model showed a good or moderate predictive ability, whereas the linear regression model using only eGFR data did not.

## Disclosures

O.K. and S.T. are employees of JMDC Inc. M.I. previously received honoraria from JMDC Inc. for conference presentations and academic consultations, but does not receive any fee for the present study.

## Funding

none (self-funded)

## Acknowledgments

We thank Editage (www.editage.com) for English language editing.

## Author contributions

M.I., O.K., and S.T. planned the study. O.K. and S.T. collected and analyzed the data. M.I. wrote the manuscript. O.K. and S.T. prepared the tables, figures, and supplementary materials. All the authors have reviewed the final version of the manuscript.

## Data availability

The data used in this study were licensed by JMDC Inc. Proposals and requests for data access should be directed to the corresponding authors via email.

## Supplementary materials

**Supplementary Table S1**. **Anatomical Therapeutic Chemical Classification codes to define each drug**

**Supplementary Table S2**. **List of predictor variables from 2012 to 2016 and distributions by the status of data availability from 2017 to 2021**

**Supplementary Figure S1. Study flow chart**

**Supplementary Table S3. The mean (standard deviation) slope of eGFR (ml/min/1.73 m^2^/year) for the 10 years by age, sex, and KDIGO GFR stages in 2016**

## References

1. Go AS, Chertow GM, Fan D, et al. Chronic kidney disease and the risks of death, cardiovascular events, and hospitalization. N Engl J Med. 2004;351(13):1296–1305. doi:10.1056/NEJMoa041031

2. Iwagami M, Caplin B, Smeeth L, et al. Chronic kidney disease and cause-specific hospitalisation: a matched cohort study using primary and secondary care patient data. Br J Gen Pract. 2018;68(673):e512–e523. doi:10.3399/bjgp18X697973

3. Sakoi N, Mori Y, Tsugawa Y, et al. Early-stage chronic kidney disease and related health care spending. JAMA Netw Open. 2024;7(1):e2351518. doi:10.1001/jamanetworkopen.2023.51518

4. Khan SS, Kazmi WH, Abichandani R, et al. Health care utilization among patients with chronic kidney disease. Kidney Int. 2002;62(1):229–236. doi:10.1046/j.1523-1755.2002.00432.x

5. Fujii M, Ohno Y, Ikeda A, et al. Current status of the rapid decline in renal function due to diabetes mellitus and its associated factors: analysis using the National Database of Health Checkups in Japan. Hypertens Res. 2023;46(5):1075–1089. doi:10.1038/s41440-023-01185-2

6. Lindeman RD, Tobin J, Shock NW. Longitudinal studies on the rate of decline in renal function with age. J Am Geriatr Soc. 1985;33(4):278–285. doi:10.1111/j.1532-5415.1985.tb07117.x

7. Cleary F, Prieto-Merino D, Nitsch D. A systematic review of statistical methodology used to evaluate progression of chronic kidney disease using electronic healthcare records. PLOS ONE. 2022;17(7):e0264167. doi:10.1371/journal.pone.0264167

8. Baigent C, Herrington WG, Coresh J, et al. Challenges in conducting clinical trials in nephrology: conclusions from a Kidney Disease-Improving Global Outcomes (KDIGO) Controversies Conference. Kidney Int. 2017;92(2):297–305. doi:10.1016/j.kint.2017.04.019

9. Carrero JJ, Fu EL, Vestergaard SV, et al. Defining measures of kidney function in observational studies using routine health care data: methodological and reporting considerations. Kidney Int. 2023;103(1):53–69. doi:10.1016/j.kint.2022.09.020

10. Itano S, Kanda E, Nagasu H, et al. eGFR slope as a surrogate endpoint for clinical study in early stage of chronic kidney disease: from The Japan Chronic Kidney Disease Database. Clin Exp Nephrol. 2023;27(10):847–856. doi:10.1007/s10157-023-02376-4

11. Levey AS, Gansevoort RT, Coresh J, et al. Change in albuminuria and GFR as end points for clinical trials in early stages of CKD: a scientific workshop sponsored by the National Kidney Foundation in collaboration with the US Food and Drug Administration and European Medicines Agency. Am J Kidney Dis. 2020;75(1):84–104. doi:10.1053/j.ajkd.2019.06.009

12. Grams ME, Sang Y, Ballew SH, et al. Evaluating glomerular filtration rate slope as a surrogate end point for ESKD in clinical trials: an individual participant meta-analysis of observational data. J Am Soc Nephrol. 2019;30(9):1746–1755. doi:10.1681/ASN.2019010008

13. Inker LA, Collier W, Greene T, et al. A meta-analysis of GFR slope as a surrogate endpoint for kidney failure. Nat Med. 2023;29(7):1867–1876. doi:10.1038/s41591-023-02418-0

14. Rifkin DE, Shlipak MG, Katz R, et al. Rapid kidney function decline and mortality risk in older adults. Arch Intern Med. 2008;168(20):2212–2218. doi:10.1001/archinte.168.20.2212

15. Tangri N, Grams ME, Levey AS, et al. Multinational assessment of accuracy of equations for predicting risk of kidney failure: a meta-analysis. JAMA. 2016;315(2):164–174. doi:10.1001/jama.2015.18202

16. Tangri N, Stevens LA, Griffith J, et al. A predictive model for progression of chronic kidney disease to kidney failure. JAMA. 2011;305(15):1553–1559. doi:10.1001/jama.2011.451

17. Tsai MK, Gao W, Chien KL, et al. A prediction model with lifestyle factors improves the predictive ability for renal replacement therapy: a cohort of 442 714 Asian adults. Clin Kidney J. 2022;15(10):1896–1907. doi:10.1093/ckj/sfac119

18. Grams ME, Brunskill NJ, Ballew SH, et al. Development and validation of prediction models of adverse kidney outcomes in the population with and without diabetes. Diabetes Care. 2022;45(9):2055–2063. doi:10.2337/dc22-0698

19. Aoki J, Kaya C, Khalid O, et al. CKD progression prediction in a diverse US population: a machine-learning model. Kidney Med. 2023;5(9):100692. doi:10.1016/j.xkme.2023.100692

20. Chan L, Nadkarni GN, Fleming F, et al. Derivation and validation of a machine learning risk score using biomarker and electronic patient data to predict progression of diabetic kidney disease. Diabetologia. 2021;64(7):1504–1515. doi:10.1007/s00125-021-05444-0

21. OECD. OECD Reviews of Public Health: Japan: A Healthier Tomorrow. OECD Publishing; 2019.

22. Ministry of Health, Labour and Welfare. Implementation status of specific health checkups and specific health guidance in 2021. https://www.mhlw.go.jp/stf/seisakunitsuite/bunya/newpage_00043.html.

23. Nagai K, Tanaka T, Kodaira N, et al. Data resource profile: JMDC claims database sourced from health insurance societies. J Gen Fam Med. 2021;22(3):118–127. doi:10.1002/jgf2.422

24. Matsuo S, Imai E, Horio M, et al. Revised equations for estimated GFR from serum creatinine in Japan. Am J Kidney Dis. 2009;53(6):982–992. doi:10.1053/j.ajkd.2008.12.034

25. Chen T, Guestrin C. Xgboost: a scalable tree boosting system. In: Proceedings of the 22nd ACM SIGKDD International Conference on Knowledge Discovery and Data Mining; 2016:785–794. doi:10.1145/2939672.2939785

26. Lundberg SM, Erion GG, Lee S-I. Consistent individualized feature attribution for tree ensembles. doi:10.48550/arXiv.1802.03888

27. Baba M, Shimbo T, Horio M, et al. Longitudinal study of the decline in renal function in healthy subjects. PLOS ONE. 2015;10(6):e0129036. doi:10.1371/journal.pone.0129036

28. Masrouri S, Alijanzadeh D, Amiri M, et al. Predictors of decline in kidney function in the general population: a decade of follow-up from the Tehran Lipid and glucose Study. Ann Med. 2023;55(1):2216020. doi:10.1080/07853890.2023.2216020

29. Jaques DA, Vollenweider P, Bochud M, et al. Aging and hypertension in kidney function decline: a 10 year population-based study. Front Cardiovasc Med. 2022;9:1035313. doi:10.3389/fcvm.2022.1035313

30. Cohen E, Nardi Y, Krause I, et al. A longitudinal assessment of the natural rate of decline in renal function with age. J Nephrol. 2014;27(6):635–641. doi:10.1007/s40620-014-0077-9

31. Tsai CW, Ting IW, Yeh HC, et al. Longitudinal change in estimated GFR among CKD patients: a 10-year follow-up study of an integrated kidney disease care program in Taiwan. PLOS ONE. 2017;12(4):e0173843. doi:10.1371/journal.pone.0173843

32. Imai E, Horio M, Yamagata K, et al. Slower decline of glomerular filtration rate in the Japanese general population: a longitudinal 10-year follow-up study. Hypertens Res. 2008;31(3):433–441. doi:10.1291/hypres.31.433

33. Hsu CY, McCulloch CE, Iribarren C, et al. Body mass index and risk for end-stage renal disease. Ann Intern Med. 2006;144(1):21–28. doi:10.7326/0003-4819-144-1-200601030-00006

34. Lew QJ, Jafar TH, Talaei M, et al. Increased body mass index is a risk factor for end-stage renal disease in the Chinese Singapore population. Kidney Int. 2017;92(4):979–987. doi:10.1016/j.kint.2017.03.019

35. Abdelhafiz AH, Tan E, Levett C, et al. Natural history and predictors of faster glomerular filtration rate decline in a referred population of older patients with type 2 diabetes mellitus. Hosp Pract (1995). 2012;40(4):49–55. doi:10.3810/hp.2012.10.1003

