## Supplementary material for "Estimating and predicting kidney function decline in the general population": Supplemantary Tables 1 to 3 and Figure 1

**Supplementary Table S1. Anatomical Therapeutic Chemical Classification codes to define each drug**

| **Drug** | **Anatomical Therapeutic Chemical Classification codes** |
|---|---|
| Lipid-lowering agents (any) | C10, C11 |
| Statins | C10A1 |
| Antidiabetic drugs (any) | A10 |
| Sodium-glucose cotransporter-2 inhibitors | A10P1 |
| Antihypertensive drugs (any) | C02, C03, C07, C08, C09, C11 |
| Angiotensin converting enzyme inhibitor or angiotensin receptor blocker | C09 |
| Antiplatelet drugs | B01 |

**Supplementary Table S2. List of predictor variables from 2012 to 2016 and distributions by the status of data availability from 2017 to 2021**

| **Variables** | **People with the annual health checkup results (including serum creatinine) for the first 5 years from 2012 to 2016 (N = 183 364)** | | | |
|---|---|---|---|---|
| | **People with the annual health checkup results (including serum creatinine) for the latter 5 years from 2017 to 2021 (N = 126 424)** | | **People without the annual health checkup results (including serum creatinine) for the latter 5 years from 2017 to 2021 (N = 56 926)** | |
| | **Distribution** | **Missing** | **Distribution** | **Missing** |
| Age (years) | Mean 45.2, SD 8.6 | 0 (0%) | Mean 48.6, SD 11.8 | 0 (0%) |
| Sex | Men: 104 219 (82.4%), Women: 22 205 (17.6%) | 0 (0%) | Men: 39 367 (69.2%), Women: 17 559 (30.9%) | 0 (0%) |
| **Results of annual health checkups in 2016** | | | | |
| eGFR (ml/min/1.73m$^2$) | Mean 79.0, SD 13.4 | 0 (0%) | Mean 78.0, SD 14.7 | 0 (0%) |
| Body mass index (kg/m$^2$) | Mean 23.4, SD 3.6 | 18 (0.01%) | Mean 23.1, SD 3.6 | 24 (0.04%) |
| Abdominal circumference (cm) | Mean 82.6, SD 9.6 | 10 435 (8.3%) | Mean 82.3, SD 9.7 | 7 575 (13.3%) |
| Systolic blood pressure (mmHg) | Mean 121.6, SD 14.4 | 20 (0.02%) | Mean 121.9, SD 15.8 | 20 (0.04%) |
| Diastolic blood pressure (mmHg) | Mean 75.6, SD 11.1 | 20 (0.02%) | Mean 75.1, SD 11.4 | 20 (0.04%) |
| Triglycerides (mg/dl) | Mean 115.7, SD 91.6 | 31 (0.02%) | Mean 110.1, SD 83.5 | 31 (0.1%) |
| High density lipoprotein cholesterol (mg/dl) | Mean 60.7, SD 16.1 | 17 (0.01%) | Mean 63.3, SD 17.0 | 27 (0.1%) |
| Low density lipoprotein cholesterol (mg/dl) | Mean 120.7, SD 29.7 | 29 (0.02%) | Mean 120.0, SD 30.4 | 32 (0.1%) |
| Total cholesterol (mg/dl) | Mean 201.8, SD 33.6 | 76 961 (60.9%) | Mean 204.4, SD 34.4 | 31 767 (55.8%) |
| Aspartate aminotransferase (U/l) | Mean 22.4, SD 10.1 | 14 (0.01%) | Mean 22.4, SD 11.1 | 18 (0.03%) |
| Alanine aminotransferase (U/l) | Mean 25.3, SD 18.8 | 14 (0.01%) | Mean 23.3, SD 17.6 | 18 (0.03%) |
| Gamma glutamyl transpeptidase (U/l) | Mean 39.8, SD 43.4 | 30 (0.02%) | Mean 38.4, SD 46.0 | 36 (0.1%) |
| Fasting blood sugar (mg/dl) | Mean 94.4, SD 16.7 | 27 532 (21.8%) | Mean 96.3, SD 18.7 | 7 882 (13.9%) |
| Casual blood sugar (mg/dl) | Mean 98.5, SD 25.9 | 116 912 (92.5%) | Mean 100.0, SD 29.9 | 51 751 (90.9%) |
| Hemoglobin A1c (NGSP) (%) | Mean 5.5, SD 0.6 | 37 179 (29.4%) | Mean 5.6, SD 0.7 | 16 951 (29.8%) |
| Hematocrit (%) | Mean 45.1, SD 3.8 | 6 895 (5.5%) | Mean 44.1, SD 4.0 | 1 423 (2.5%) |

| | | | | |
|---|---|---|---|---|
| Hemoglobin (g/dl) | Mean 14.8, SD 1.3 | 790 (0.6%) | Mean 14.5, SD 1.5 | 332 (0.6%) |
| Red blood cells ($10^6$/µl) | Mean 488.1, SD 41.1 | 575 (0.5%) | Mean 478.2, SD 44.5 | 268 (0.5%) |
| Serum uric acid (mg/dl) | Mean 5.7, SD 1.3 | 6 429 (5.1%) | Mean 5.5, SD 1.4 | 2 827 (5.0%) |
| Urinary sugar (dipstick test) | -: 120 612 (95.4%), +/-: 628 (0.5%), +: 738 (0.6%), ++: 477 (0.4%), +++: 906 (0.7%) | 3 063 (2.4%) | -: 54 629 (96.0%), +/-: 299 (0.5%), +: 364 (0.6%), ++: 302 (0.5%), +++: 496 (0.9%) | 836 (1.5%) |
| Urinary protein (dipstick test) | -: 112 157 (88.7%), +/-: 7 961 (6.3%), +: 2 437 (1.9%), ++: 656 (0.5%), +++: 166 (0.1%) | 3 047 (2.4%) | -: 50 443 (88.6%), +/-: 3 918 (6.9%), +: 1 195 (2.1%), ++: 440 (0.8%), +++: 111 (0.2%) | 819 (1.4%) |
| Smoking status | Yes: 36 760 (29.1%), No: 81 194 (64.2%) | 8 470 (6.7%) | Yes: 13 758 (24.2%), No: 40 282 (70.8%) | 2 886 (5.1%) |
| Drinking habits | Every day: 31 809 (25.2%), Sometimes: 37 867 (30.0%), Rarely/none: 47 576 (37.6%) | 9 172 (7.3%) | Every day: 13 927 (24.5%), Sometimes: 16 133 (28.3%), Rarely/none: 23 024 (40.5%) | 3 842 (6.8%) |
| Exercise (≥2/week and ≥30 minutes in the past year) | Yes: 22 549 (17.8%), No: 86 889 (68.7%) | 16 986 (13.4%) | Yes: 12 117 (21.3%), No: 39 685 (69.7%) | 5 124 (9.0%) |
| **Prescriptions in 2016** | | | | |
| Lipid-lowering agents (any) | Yes: 12 979 (10.3%), No: 113 445 (89.7%) | 0 (0%) | Yes: 8 176 (14.4%), No: 48 750 (85.6%) | 0 (0%) |
| Statins | Yes: 10 262 (8.1%), No: 116 162 (91.9%) | 0 (0%) | Yes: 6 619 (11.6%), No: 50 307 (88.4%) | 0 (0%) |
| Antidiabetic drugs (any) | Yes: 4 428 (3.5%), No: 121 996 (96.5%) | 0 (0%) | Yes: 3 077 (5.4%), No: 53 849 (94.6%) | 0 (0%) |
| Sodium-glucose cotransporter-2 inhibitors | Yes: 1 106 (0.9%), No: 125 318 (99.1%) | 0 (0%) | Yes: 565 (1.0%), No: 56 361 (99.0%) | 0 (0%) |
| Antihypertensive drugs (any) | Yes: 14 885 (11.8%), No: 111 539 (88.2%) | 0 (0%) | Yes: 9 939 (17.5%), No: 46 987 (82.5%) | 0 (0%) |

| | | | | |
|---|---|---|---|---|
| ACEI or ARB | Yes: 10 038 (7.9%), No: 116 386 (92.1%) | 0 (0%) | Yes: 6 610 (11.6%), No: 50 316 (88.4%) | 0 (0%) |
| Antiplatelet drugs | Yes: 2 948 (2.3%), No: 123 476 (97.7%) | 0 (0%) | Yes: 2 470 (4.3%), No: 54 456 (95.7%) | 0 (0%) |
| **Results of annual health checkups in 2015** | | | | |
| eGFR (ml/min/1.73m$^2$) | Mean 80.3, SD 13.5 | 0 (0%) | Mean 78.9, SD 14.7 | 0 (0%) |
| Body mass index (kg/m$^2$) | Mean 23.3, SD 3.6 | 17 (0.01%) | Mean 23.0, SD 3.6 | 7 (0.01%) |
| Abdominal circumference (cm) | Mean 82.3, SD 9.6 | 11 784 (9.3%) | Mean 82.1, SD 9.7 | 7 836 (13.8%) |
| Systolic blood pressure (mmHg) | Mean 121.2, SD 14.3 | 12 (0.01%) | Mean 121.4, SD 15.6 | 9 (0.02%) |
| Diastolic blood pressure (mmHg) | Mean 75.1, SD 11.0 | 12 (0.01%) | Mean 74.7, SD 11.4 | 9 (0.02%) |
| Triglycerides (mg/dl) | Mean 114.4, SD 88.7 | 113 (0.1%) | Mean 109.3, SD 86.8 | 81 (0.1%) |
| High density lipoprotein cholesterol (mg/dl) | Mean 60.7, SD 16.1 | 18 (0.01%) | Mean 63.1, SD 16.9 | 17 (0.03%) |
| Low density lipoprotein cholesterol (mg/dl) | Mean 120.4, SD 30.1 | 22 (0.02%) | Mean 119.7, SD 30.7 | 19 (0.03%) |
| Total cholesterol (mg/dl) | Mean 201.2, SD 33.7 | 76 955 (60.9%) | Mean 204.1, SD 34.5 | 31 883 (56.0%) |
| Aspartate aminotransferase (U/l) | Mean 22.3, SD 10.5 | 13 (0.01%) | Mean 22.2, SD 11.3 | 5 (0.01%) |
| Alanine aminotransferase (U/l) | Mean 25.0, SD 18.7 | 14 (0.01%) | Mean 23.0, SD 17.4 | 5 (0.01%) |
| Gamma glutamyl transpeptidase (U/l) | Mean 39.0, SD 42.8 | 28 (0.02%) | Mean 37.8, SD 46.1 | 13 (0.02%) |
| Fasting blood sugar (mg/dl) | Mean 93.9, SD 16.6 | 28 416 (22.5%) | Mean 95.8, SD 18.1 | 8 169 (14.4%) |
| Casual blood sugar (mg/dl) | Mean 96.7, SD 22.3 | 111 655 (88.3%) | Mean 98.1, SD 24.7 | 49 606 (87.1%) |
| Hemoglobin A1c (NGSP) (%) | Mean 5.5, SD 0.6 | 37 914 (30.0%) | Mean 5.6, SD 0.7 | 17 226 (30.3%) |
| Hematocrit (%) | Mean 44.8, SD 3.7 | 6 725 (5.3%) | Mean 43.9, SD 3.9 | 1 335 (2.4%) |
| Hemoglobin (g/dl) | Mean 14.8, SD 1.3 | 854 (0.7%) | Mean 14.5, SD 1.5 | 323 (0.6%) |
| Red blood cells ($10^6$/μl) | Mean 487.3, SD 40.2 | 650 (0.5%) | Mean 477.6, SD 43.2 | 266 (0.5%) |
| Serum uric acid (mg/dl) | Mean 5.7, SD 1.3 | 6 169 (4.9%) | Mean 5.5, SD 1.4 | 2 627 (4.6%) |
| Urinary sugar (dipstick test) | -: 120 725 (95.5%), +/-: 572 (0.5%), +: 696 (0.6%), ++: 460 (0.4%), +++: 580 | 3 391 (2.7%) | -: 54 795 (96.3%), +/-: 262 (0.5%), +: 352 (0.6%), ++: 265 (0.5%), +++: 338 | 914 (1.6%) |

| | | | | |
|---|---|---|---|---|
| | (0.5%) | | (0.6%) | |
| Urinary protein (dipstick test) | -: 112 527 (89.0%), +/-: 7 488 (5.9%), +: 2 239 (1.8%), ++: 632 (0.5%), +++: 157 (0.1%) | 3 381 (2.7%) | -: 50 824 (89.3%), +/-: 3 532 (6.2%), +: 1 138 (2.0%), ++: 412 (0.7%), +++: 111 (0.2%) | 909 (1.6%) |
| Smoking status | Yes: 36 852 (29.2%), No: 80 155 (63.4%) | 9 417 (7.5%) | Yes: 13 936 (24.5%), No: 39 904 (70.1%) | 3 086 (5.4%) |
| Drinking habits | Every day: 31 681 (25.1%), Sometimes: 37 583 (29.7%), Rarely/none: 46 447 (36.7%) | 10 713 (8.5%) | Every day: 14 001 (24.6%), Sometimes: 16 266 (28.6%), Rarely/none: 22 487 (39.5%) | 4 172 (7.3%) |
| Exercise (≥2/week and ≥30 minutes in the past year) | Yes: 21 683 (17.2%), No: 86 077 (68.1%) | 18 664 (14.8%) | Yes: 11 683 (20.5%), No: 39 644 (69.6%) | 5 599 (9.8%) |
| **Prescriptions in 2015** | | | | |
| Lipid-lowering agents (any) | Yes: 12 126 (9.6%), No: 114 298 (90.4%) | 0 (0%) | Yes: 7 700 (13.5%), No: 49 226 (86.5%) | 0 (0%) |
| Statins | Yes: 9 562 (7.6%), No: 116 862 (92.4%) | 0 (0%) | Yes: 6 244 (11.0%), No: 50 682 (89.0%) | 0 (0%) |
| Antidiabetic drugs (any) | Yes: 3 994 (3.2%), No: 122 430 (96.8%) | 0 (0%) | Yes: 2 844 (5.0%), No: 54 082 (95.0%) | 0 (0%) |
| Sodium-glucose cotransporter-2 inhibitors | Yes: 652 (0.5%), No: 125 772 (99.5%) | 0 (0%) | Yes: 344 (0.6%), No: 56 582 (99.4%) | 0 (0%) |
| Antihypertensive drugs (any) | Yes: 13 581 (10.7%), No: 112 843 (89.3%) | 0 (0%) | Yes: 9 172 (16.1%), No: 47 754 (83.9%) | 0 (0%) |
| ACEI or ARB | Yes: 9 212 (7.3%), No: 117 212 (92.7%) | 0 (0%) | Yes: 6 166 (10.8%), No: 50 760 (89.2%) | 0 (0%) |
| Antiplatelet drugs | Yes: 2 808 (2.2%), No: 123 616 (97.8%) | 0 (0%) | Yes: 2 206 (3.9%), No: 54 720 (96.1%) | 0 (0%) |
| **Results of annual health checkups in 2014** | | | | |

| | | | | |
|---|---|---|---|---|
| eGFR (ml/min/1.73m$^2$) | Mean 81.6, SD 13.7 | 0 (0%) | Mean 80.2, SD 15.0 | 0 (0%) |
| Body mass index (kg/m$^2$) | Mean 23.3, SD 3.6 | 19 (0.02%) | Mean 22.9, SD 3.6 | 8 (0.01%) |
| Abdominal circumference (cm) | Mean 82.1, SD 9.5 | 13 300 (10.5%) | Mean 81.8, SD 9.6 | 8 053 (14.2%) |
| Systolic blood pressure (mmHg) | Mean 121.0, SD 14.2 | 18 (0.01%) | Mean 121.0, SD 15.5 | 6 (0.01%) |
| Diastolic blood pressure (mmHg) | Mean 74.8, SD 10.9 | 18 (0.01%) | Mean 74.4, SD 11.3 | 6 (0.01%) |
| Triglycerides (mg/dl) | Mean 114.1, SD 88.9 | 520 (0.4%) | Mean 108.2, SD 84.0 | 215 (0.4%) |
| High density lipoprotein cholesterol (mg/dl) | Mean 60.8, SD 15.9 | 27 (0.02%) | Mean 63.2, SD 16.7 | 9 (0.02%) |
| Low density lipoprotein cholesterol (mg/dl) | Mean 120.1, SD 30.1 | 30 (0.02%) | Mean 119.5, SD 30.7 | 13 (0.02%) |
| Total cholesterol (mg/dl) | Mean 199.7, SD 33.3 | 76 679 (60.7%) | Mean 202.7, SD 34.3 | 31 776 (55.8%) |
| Aspartate aminotransferase (U/l) | Mean 22.0, SD 10.3 | 14 (0.01%) | Mean 21.9, SD 10.4 | 1 (0.002%) |
| Alanine aminotransferase (U/l) | Mean 25.0, SD 19.3 | 15 (0.01%) | Mean 22.9, SD 17.0 | 1 (0.002%) |
| Gamma glutamyl transpeptidase (U/l) | Mean 38.1, SD 41.3 | 22 (0.02%) | Mean 36.8, SD 43.7 | 18 (0.03%) |
| Fasting blood sugar (mg/dl) | Mean 93.2, SD 16.6 | 28 453 (22.5%) | Mean 95.3, SD 18.4 | 8 179 (14.4%) |
| Casual blood sugar (mg/dl) | Mean 96.4, SD 21.8 | 111 352 (88.1%) | Mean 98.0, SD 25.3 | 49 513 (87.0%) |
| Hemoglobin A1c (NGSP) (%) | Mean 5.5, SD 0.6 | 37 912 (30.0%) | Mean 5.6, SD 0.6 | 17 450 (30.7%) |
| Hematocrit (%) | Mean 44.7, SD 3.7 | 6 669 (5.3%) | Mean 43.8, SD 4.0 | 1 250 (2.2%) |
| Hemoglobin (g/dl) | Mean 14.8, SD 1.4 | 716 (0.6%) | Mean 14.5, SD 1.5 | 221 (0.4%) |
| Red blood cells (10$^6$/μl) | Mean 486.4, SD 40.3 | 542 (0.4%) | Mean 477.1, SD 43.2 | 183 (0.3%) |
| Serum uric acid (mg/dl) | Mean 5.7, SD 1.3 | 5 877 (4.7%) | Mean 5.5, SD 1.4 | 2 355 (4.1%) |
| Urinary sugar (dipstick test) | -: 119 976 (94.9%), +/-: 604 (0.5%), +: 740 (0.6%), ++: 402 (0.3%), +++: 435 (0.3%) | 4 267 (3.4%) | -: 54 527 (95.8%), +/-: 286 (0.5%), +: 349 (0.6%), ++: 257 (0.5%), +++: 274 (0.5%) | 1 233 (2.2%) |
| Urinary protein (dipstick test) | -: 110 475 (87.4%), +/-: 8 440 (6.7%), +: 2 478 (2.0%), ++: 610 (0.5%), +++: 135 (0.1%) | 4 286 (3.4%) | -: 49 664 (87.2%), +/-: 4 253 (7.5%), +: 1 250 (2.2%), ++: 396 (0.7%), +++: 109 (0.2%) | 1 254 (2.2%) |
| Smoking status | Yes: 37 275 (29.5%), No: 78 734 | 10 415 (8.2%) | Yes: 14 217 (25.0%), No: 39 378 | 3 331 (5.9%) |

| | | | | |
|---|---|---|---|---|
| | (62.3%) | | (69.2%) | |
| Drinking habits | Every day: 31 036 (24.6%), Sometimes: 37 515 (29.7%), Rarely/none: 46 230 (36.6%) | 11 643 (9.2%) | Every day: 13 753 (24.2%), Sometimes: 16 350 (28.7%), Rarely/none: 22 275 (39.1%) | 4 548 (8.0%) |
| Exercise (≥2/week and ≥30 minutes in the past year) | Yes: 20 893 (16.5%), No: 86 372 (68.3%) | 19 159 (15.2%) | Yes: 11 338 (19.9%), No: 39 778 (69.9%) | 5 810 (10.2%) |
| **Prescriptions in 2014** | | | | |
| Lipid-lowering agents (any) | Yes: 11 312 (9.0%), No: 115 112 (91.1%) | 0 (0%) | Yes: 7 246 (12.7%), No: 49 680 (87.3%) | 0 (0%) |
| Statins | Yes: 8 902 (7.0%), No: 117 522 (93.0%) | 0 (0%) | Yes: 5 869 (10.3%), No: 51 057 (89.7%) | 0 (0%) |
| Antidiabetic drugs (any) | Yes: 3 557 (2.8%), No: 122 867 (97.2%) | 0 (0%) | Yes: 2 655 (4.7%), No: 54 271 (95.3%) | 0 (0%) |
| Sodium-glucose cotransporter-2 inhibitors | Yes: 225 (0.2%), No: 126 199 (99.8%) | 0 (0%) | Yes: 149 (0.3%), No: 56 777 (99.7%) | 0 (0%) |
| Antihypertensive drugs (any) | Yes: 12 410 (9.8%), No: 114 014 (90.2%) | 0 (0%) | Yes: 8 581 (15.1%), No: 48 345 (84.9%) | 0 (0%) |
| ACEI or ARB | Yes: 8 500 (6.7%), No: 117 924 (93.3%) | 0 (0%) | Yes: 5 766 (10.1%), No: 51 160 (89.9%) | 0 (0%) |
| Antiplatelet drugs | Yes: 2 567 (2.0%), No: 123 857 (98.0%) | 0 (0%) | Yes: 2 005 (3.5%), No: 54 921 (96.5%) | 0 (0%) |
| **Results of annual health checkups in 2013** | | | | |
| eGFR (ml/min/1.73m$^2$) | Mean 82.0, SD 13.9 | 0 (0%) | Mean 80.8, SD 15.1 | 0 (0%) |
| Body mass index (kg/m$^2$) | Mean 23.2, SD 3.6 | 23 (0.02%) | Mean 22.9, SD 3.5 | 44 (0.1%) |
| Abdominal circumference (cm) | Mean 81.9, SD 9.5 | 16 434 (13.0%) | Mean 81.7, SD 9.5 | 8 594 (15.1%) |
| Systolic blood pressure (mmHg) | Mean 120.8, SD 14.2 | 21 (0.02%) | Mean 120.8, SD 15.4 | 40 (0.1%) |
| Diastolic blood pressure (mmHg) | Mean 74.5, SD 10.9 | 21 (0.02%) | Mean 74.2, SD 11.3 | 40 (0.1%) |

| | | | | |
|---|---|---|---|---|
| Triglycerides (mg/dl) | Mean 113.8, SD 88.6 | 527 (0.4%) | Mean 108.5, SD 85.0 | 246 (0.4%) |
| High density lipoprotein cholesterol (mg/dl) | Mean 60.2, SD 15.6 | 30 (0.02%) | Mean 62.3, SD 16.5 | 40 (0.1%) |
| Low density lipoprotein cholesterol (mg/dl) | Mean 118.6, SD 30.1 | 35 (0.03%) | Mean 118.1, SD 30.7 | 43 (0.1%) |
| Total cholesterol (mg/dl) | Mean 199.2, SD 33.6 | 83 023 (65.7%) | Mean 202.1, SD 34.7 | 34 701 (61.0%) |
| Aspartate aminotransferase (U/l) | Mean 21.9, SD 10.1 | 10 (0.01%) | Mean 21.8, SD 10.1 | 33 (0.1%) |
| Alanine aminotransferase (U/l) | Mean 25.0, SD 19.0 | 11 (0.01%) | Mean 22.9, SD 17.2 | 33 (0.1%) |
| Gamma glutamyl transpeptidase (U/l) | Mean 37.7, SD 41.0 | 17 (0.01%) | Mean 36.7, SD 43.6 | 37 (0.1%) |
| Fasting blood sugar (mg/dl) | Mean 93.0, SD 16.9 | 25 009 (19.8%) | Mean 95.1, SD 19.1 | 6 841 (12.0%) |
| Casual blood sugar (mg/dl) | Mean 96.1, SD 22.4 | 111 921 (88.5%) | Mean 97.7, SD 26.3 | 49 954 (87.8%) |
| Hemoglobin A1c (NGSP) (%) | Mean 5.5, SD 0.6 | 38 181 (30.2%) | Mean 5.6, SD 0.7 | 17 683 (31.1%) |
| Hematocrit (%) | Mean 44.9, SD 3.8 | 6 818 (5.4%) | Mean 44.0, SD 4.0 | 1 380 (2.4%) |
| Hemoglobin (g/dl) | Mean 14.9, SD 1.4 | 840 (0.7%) | Mean 14.6, SD 1.5 | 349 (0.6%) |
| Red blood cells ($10^6$/μl) | Mean 487.3, SD 40.8 | 704 (0.6%) | Mean 477.8, SD 43.4 | 301 (0.5%) |
| Serum uric acid (mg/dl) | Mean 5.6, SD 1.3 | 22 535 (17.8%) | Mean 5.4, SD 1.4 | 8 235 (14.5%) |
| Urinary sugar (dipstick test) | -: 120 795 (95.6%), +/-: 607 (0.5%), +: 721 (0.6%), ++: 381 (0.3%), +++: 386 (0.3%) | 3 534 (2.8%) | -: 54 570 (95.9%), +/-: 298 (0.5%), +: 364 (0.6%), ++: 239 (0.4%), +++: 234 (0.4%) | 1 221 (2.1%) |
| Urinary protein (dipstick test) | -: 111 202 (88.0%), +/-: 8 282 (6.6%), +: 2 666 (2.1%), ++: 604 (0.5%), +++: 141 (0.1%) | 3 529 (2.8%) | -: 49 847 (87.6%), +/-: 4 131 (7.3%), +: 1 317 (2.3%), ++: 338 (0.6%), +++: 105 (0.2%) | 1 188 (2.1%) |
| Smoking status | Yes: 37 797 (29.9%), No: 77 016 (60.9%) | 11 611 (9.2%) | Yes: 14 589 (25.6%), No: 38 693 (68.0%) | 3 644 (6.4%) |
| Drinking habits | Every day: 30 487 (24.1%), Sometimes: 36 935 (29.2%), Rarely/none: 45 664 (36.1%) | 13 338 (10.6%) | Every day: 13 612 (23.9%), Sometimes: 16 252 (28.6%), Rarely/none: 22 056 (38.8%) | 5 006 (8.8%) |
| Exercise (≥2/week and ≥30 minutes in the | Yes: 19 992 (15.8%), No: 85 555 | 20 877 (16.5%) | Yes: 10 795 (19.0%), No: 39 821 | 6 310 (11.1%) |

| | | | | |
|---|---|---|---|---|
| past year) | (67.7%) | | (70.0%) | |
| **Prescriptions in 2013** | | | | |
| Lipid-lowering agents (any) | Yes: 10 573 (8.4%), No: 115 851 (91.6%) | 0 (0%) | Yes: 6 845 (12.0%), No: 50 081 (88.0%) | 0 (0%) |
| Statins | Yes: 8 278 (6.6%), No: 118 146 (93.5%) | 0 (0%) | Yes: 5 516 (9.7%), No: 51 410 (90.3%) | 0 (0%) |
| Antidiabetic drugs (any) | Yes: 3 189 (2.5%), No: 123 235 (97.5%) | 0 (0%) | Yes: 2 405 (4.2%), No: 54 521 (95.8%) | 0 (0%) |
| Sodium-glucose cotransporter-2 inhibitors | No: 126 424 (100.0%) | 0 (0%) | No: 56 926 (100.0%) | 0 (0%) |
| Antihypertensive drugs (any) | Yes: 11 391 (9.0%), No: 115 033 (91.0%) | 0 (0%) | Yes: 8 052 (14.1%), No: 48 874 (85.9%) | 0 (0%) |
| ACEI or ARB | Yes: 7 806 (6.2%), No: 118 618 (93.8%) | 0 (0%) | Yes: 5 466 (9.6%), No: 51 460 (90.4%) | 0 (0%) |
| Antiplatelet drugs | Yes: 2 424 (1.9%), No: 124 000 (98.1%) | 0 (0%) | Yes: 1 852 (3.3%), No: 55 074 (96.8%) | 0 (0%) |
| **Results of annual health checkups in 2012** | | | | |
| eGFR (ml/min/1.73m$^2$) | Mean 82.4, SD 14.3 | 0 (0%) | Mean 80.8, SD 15.1 | 0 (0%) |
| Body mass index (kg/m$^2$) | Mean 23.1, SD 3.5 | 13 (0.01%) | Mean 22.8, SD 3.5 | 33 (0.1%) |
| Abdominal circumference (cm) | Mean 81.8, SD 9.5 | 19 936 (15.8%) | Mean 81.7, SD 9.5 | 9 196 (16.2%) |
| Systolic blood pressure (mmHg) | Mean 120.7, SD 14.1 | 9 (0.01%) | Mean 120.6, SD 15.4 | 30 (0.1%) |
| Diastolic blood pressure (mmHg) | Mean 74.2, SD 10.8 | 9 (0.01%) | Mean 74.0, SD 11.4 | 30 (0.1%) |
| Triglycerides (mg/dl) | Mean 113.2, SD 87.0 | 969 (0.8%) | Mean 107.5, SD 82.2 | 278 (0.5%) |
| High density lipoprotein cholesterol (mg/dl) | Mean 60.4, SD 15.6 | 724 (0.6%) | Mean 62.5, SD 16.3 | 221 (0.4%) |
| Low density lipoprotein cholesterol (mg/dl) | Mean 118.3, SD 30.2 | 731 (0.6%) | Mean 118.1, SD 30.9 | 227 (0.4%) |
| Total cholesterol (mg/dl) | Mean 198.7, SD 33.6 | 89 514 (70.8%) | Mean 200.8, SD 34.9 | 36 789 (64.6%) |
| Aspartate aminotransferase (U/l) | Mean 21.9, SD 11.0 | 5 (0.004%) | Mean 21.7, SD 11.3 | 27 (0.1%) |

| | | | | |
|---|---|---|---|---|
| Alanine aminotransferase (U/l) | Mean 25.0, SD 19.5 | 6 (0.005%) | Mean 22.9, SD 17.6 | 29 (0.1%) |
| Gamma glutamyl transpeptidase (U/l) | Mean 37.4, SD 40.5 | 12 (0.01%) | Mean 36.6, SD 44.4 | 36 (0.1%) |
| Fasting blood sugar (mg/dl) | Mean 92.6, SD 16.9 | 14 956 (11.8%) | Mean 94.4, SD 18.9 | 4 593 (8.1%) |
| Casual blood sugar (mg/dl) | Mean 94.8, SD 23.1 | 117 465 (92.9%) | Mean 94.7, SD 23.9 | 52 191 (91.7%) |
| Hemoglobin A1c (NGSP) (%) | Mean 5.4, SD 0.6 | 41 942 (33.2%) | Mean 5.5, SD 0.7 | 20 479 (36.0%) |
| Hematocrit (%) | Mean 45.0, SD 3.8 | 6 195 (4.9%) | Mean 44.1, SD 4.2 | 1 136 (2.0%) |
| Hemoglobin (g/dl) | Mean 14.8, SD 1.4 | 234 (0.2%) | Mean 14.5, SD 1.5 | 103 (0.2%) |
| Red blood cells ($10^6$/μl) | Mean 487.8, SD 40.3 | 55 (0.04%) | Mean 478.0, SD 43.2 | 56 (0.1%) |
| Serum uric acid (mg/dl) | Mean 5.6, SD 1.4 | 25 669 (20.3%) | Mean 5.4, SD 1.4 | 10 836 (19.0%) |
| Urinary sugar (dipstick test) | -: 119 282 (94.4%), +/-: 567 (0.5%), +: 659 (0.5%), ++: 371 (0.3%), +++: 364 (0.3%) | 5 181 (4.1%) | -: 54 319 (95.4%), +/-: 321 (0.6%), +: 358 (0.6%), ++: 222 (0.4%), +++: 235 (0.4%) | 1 471 (2.6%) |
| Urinary protein (dipstick test) | -: 110 283 (87.2%), +/-: 7 857 (6.2%), +: 2 408 (1.9%), ++: 571 (0.5%), +++: 125 (0.1%) | 5 180 (4.1%) | -: 49 760 (87.4%), +/-: 4 056 (7.1%), +: 1 208 (2.1%), ++: 362 (0.6%), +++: 92 (0.2%) | 1 448 (2.5%) |
| Smoking status | Yes: 37 908 (30.0%), No: 75 713 (59.9%) | 12 803 (10.1%) | Yes: 14 788 (26.0%), No: 38 176 (67.1%) | 3 962 (7.0%) |
| Drinking habits | Every day: 30 076 (23.8%), Sometimes: 37 057 (29.3%), Rarely/none: 44 043 (34.8%) | 15 248 (12.1%) | Every day: 13 445 (23.6%), Sometimes: 16 379 (28.8%), Rarely/none: 21 607 (38.0%) | 5 495 (9.7%) |
| Exercise (≥2/week and ≥30 minutes in the past year) | Yes: 19 101 (15.1%), No: 83 716 (66.2%) | 23 607 (18.7%) | Yes: 10 384 (18.2%), No: 39 540 (69.5%) | 7 002 (12.3%) |
| **Prescriptions in 2012** | | | | |
| Lipid-lowering agents (any) | Yes: 9 672 (7.7%), No: 116 752 (92.4%) | 0 (0%) | Yes: 6 233 (11.0%), No: 50 693 (89.1%) | 0 (0%) |
| Statins | Yes: 7 576 (6.0%), No: 118 848 (94.0%) | 0 (0%) | Yes: 5 050 (8.9%), No: 51 876 | 0 (0%) |

| | | | | |
|---|---|---|---|---|
| | | | (91.1%) | |
| Antidiabetic drugs (any) | Yes: 2 821 (2.2%), No: 123 603 (97.8%) | 0 (0%) | Yes: 2 185 (3.8%), No: 54 741 (96.2%) | 0 (0%) |
| Sodium-glucose cotransporter-2 inhibitors | No: 126 424 (100.0%) | 0 (0%) | No: 56 926 (100.0%) | 0 (0%) |
| Antihypertensive drugs (any) | Yes: 10 245 (8.1%), No: 116 179 (91.9%) | 0 (0%) | Yes: 7 406 (13.0%), No: 49 520 (87.0%) | 0 (0%) |
| ACEI or ARB | Yes: 6 943 (5.5%), No: 119 481 (94.5%) | 0 (0%) | Yes: 5 021 (8.8%), No: 51 905 (91.2%) | 0 (0%) |
| Antiplatelet drugs | Yes: 2 248 (1.8%), No: 124 176 (98.2%) | 0 (0%) | Yes: 1 744 (3.1%), No: 55 182 (96.9%) | 0 (0%) |

SD=standard deviation, eGFR=estimated glomerular filtration rate, NGSP=National Glycohemoglobin Standardization Program, ACEI=angiotensin converting enzyme inhibitor, ARB=angiotensin receptor blocker.

**Supplementary Figure S1. Study flow chart**

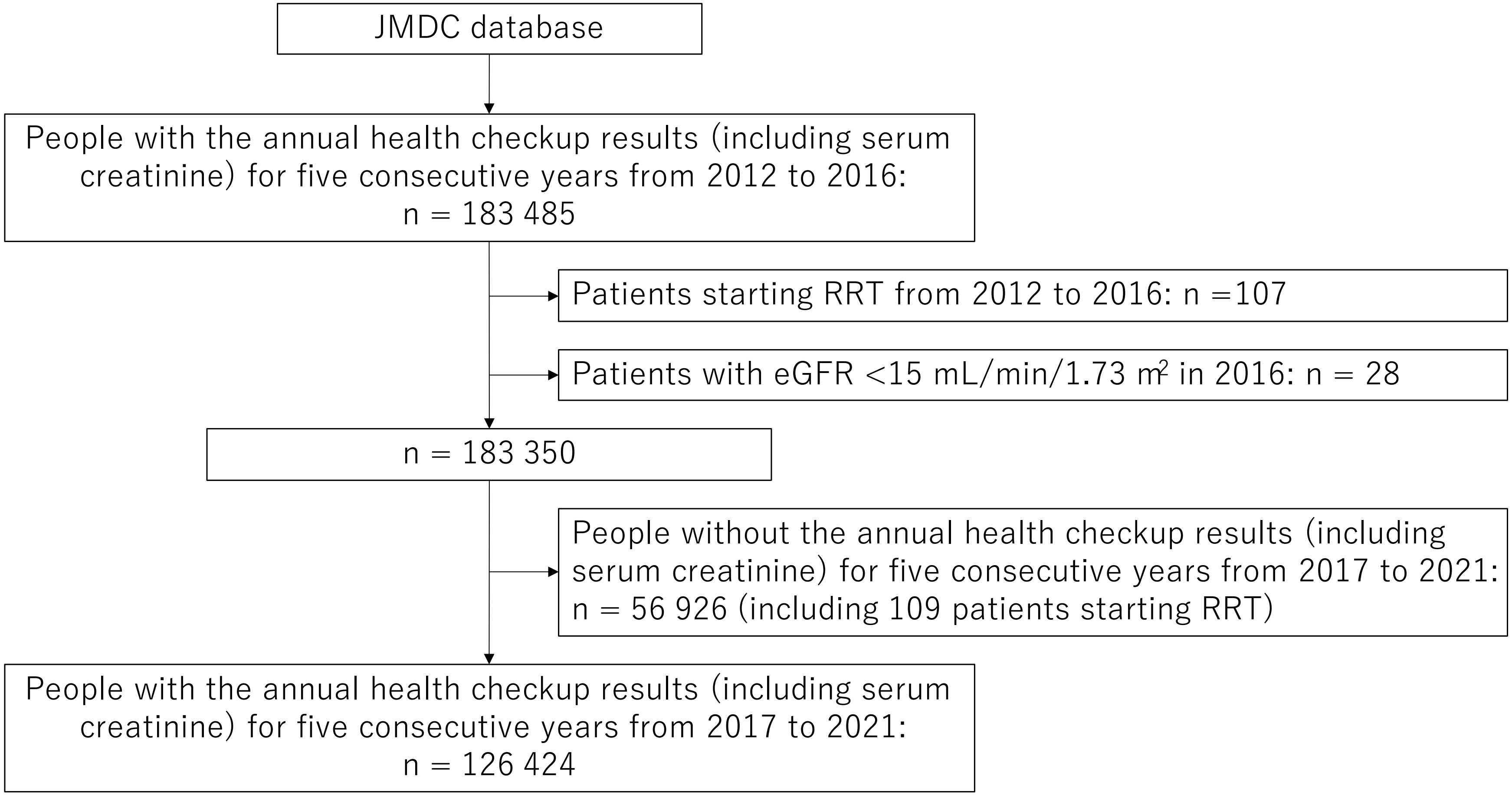


RRT, renal replacement therapy; eGFR = estimated glomerular filtration rate.

**Supplementary Table S3. The mean (standard deviation) slope of eGFR (ml/min/1.73 m2/year) for the 10 years by age, sex, and KDIGO GFR stages in 2016**

| **Men:** | **<40 years** | **40–49 years** | **50–59 years** | **≥60 years** |
|---|---|---|---|---|
| **KDIGO stage G1: eGFR ≥90 ml/min/1.73 $m^2$/year in 2016** | n = 9 776<br>Mean -1.19, SD 1.14 | n = 6 458<br>Mean -1.03, SD 1.22 | n = 3 387<br>Mean -0.80, SD 1.23 | n = 156<br>Mean -0.63, SD 1.21 |
| **KDIGO stage G2: eGFR 60–89 ml/min/1.73 $m^2$/year in 2016** | n = 18 540<br>Mean -0.99, SD 0.88 | n = 32 673<br>Mean -0.89, SD 0.86 | n = 25 189<br>Mean -0.69, SD 0.89 | n = 2 244<br>Mean -0.63, SD 0.91 |
| **KDIGO stage G3a: eGFR 45–59 ml/min/1.73 $m^2$/year in 2016** | n = 165<br>Mean -0.99, SD 1.06 | n = 1 809<br>Mean -0.85, SD 0.87 | n = 2 972<br>Mean -0.73, SD 0.90 | n = 607<br>Mean -0.64, SD 0.87 |
| **KDIGO stage G3b: eGFR 30–44 ml/min/1.73 $m^2$/year in 2016** | n = 6<br>Mean -0.70, SD 1.67 | n = 40<br>Mean -1.22, SD 1.40 | n = 128<br>Mean -1.33, SD 1.53 | n = 45<br>Mean -0.93, SD 1.17 |
| **KDIGO stage G4: eGFR 15–29 ml/min/1.73 $m^2$/year in 2016** | n = 1<br>Mean -0.84, SD n/a | n = 12<br>Mean -1.07, SD 1.24 | n = 10<br>Mean -1.47, SD 1.38 | n = 1<br>Mean -1.56, SD n/a |
| **Women:** | **<40 years** | **40–49 years** | **50–59 years** | **≥60 years** |
| **KDIGO stage G1: eGFR ≥90 ml/min/1.73 $m^2$/year in 2016** | n = 1 499<br>Mean -1.20, SD 1.34 | n = 1 927<br>Mean -1.18, SD 1.28 | n = 594<br>Mean -1.05, SD 1.32 | n = 53<br>Mean -0.93, SD 1.20 |
| **KDIGO stage G2: eGFR 60–89 ml/min/1.73 $m^2$/year in 2016** | n = 2 208<br>Mean -1.00, SD 1.04 | n = 8 881<br>Mean -0.89, SD 0.92 | n = 5 274<br>Mean -0.78, SD 0.89 | n = 507<br>Mean -0.71, SD 0.91 |
| **KDIGO stage G3a: eGFR 45–59 ml/min/1.73 $m^2$/year in 2016** | n = 23<br>Mean -0.73, SD 0.65 | n = 390<br>Mean -0.74, SD 0.67 | n = 724<br>Mean -0.70, SD 0.75 | n = 91<br>Mean -0.70, SD 0.77 |
| **KDIGO stage G3b: eGFR 30–44 ml/min/1.73 $m^2$/year in 2016** | n = 1<br>Mean -0.29, SD n/a | n = 6<br>Mean -1.33, SD 1.19 | n = 16<br>Mean -1.27, SD 1.25 | n = 5<br>Mean -1.03, SD 0.30 |
| **KDIGO stage G4: eGFR 15–29 ml/min/1.73 $m^2$/year in 2016** | n = 1<br>Mean -0.43, SD n/a | n = 2<br>Mean -1.73, SD 2.11 | n = 3<br>Mean -2.26, SD 1.01 | n = 0<br>Mean n/a, SD n/a |

KDIGO, Kidney Disease Improving Global Outcomes; eGFR, estimated glomerular filtration rate; SD, standard deviation.
